# Early Cancer Detection in Hereditary Breast and Ovarian Cancer Syndrome with Cell-Free DNA

**DOI:** 10.64898/2025.12.03.25340892

**Authors:** Erik Ensminger, Ping Luo, Julia A Sobotka, Derek Wong, Stephanie D Prokopec, Jeffrey P Bruce, Arnavaz Danesh, Jenna Eagles, Leslie Oldfield, Stephanie Pederson, Kirsten M Farncombe, Helia Purnaghshband, Adriana Aguilar-Mahecha, Ankita Nand, Beatriz E Lujan Toro, Lawrence E Heisler, Bernard Lam, Patrick Veit-Haibach, Mathieu Lupien, Mark Basik, Raymond H Kim, Trevor Pugh, CHARM Consortium

## Abstract

Early cancer detection for individuals with Hereditary Breast and Ovarian Cancer syndrome (HBOC) remains limited by the low sensitivity of available tests and lack of clinical surveillance methods for many cancers beyond breast cancer. To investigate cell-free DNA (cfDNA) sequencing as a pan-cancer surveillance modality, we analyzed 194 blood plasma samples from 88 *BRCA1* and/or *BRCA2* pathogenic variant carriers (*BRCA1/2-carriers*) using a multimodal assay integrating genomic and epigenomic (fragmentomic & DNA methylation) features. Cancer-associated signals were detected in 71% (43/61) of carriers with active cancers detected by conventional surveillance, as well as 30/54 patients (56%) with negative surveillance findings. Of the negative patients 43% (13/30) subsequently developed cancer within 2 years (12 non-breast cancers), suggesting early detection of occult cancers. These findings demonstrate the value of integrating multiple cfDNA analyses and support the potential of longitudinal, multimodal liquid biopsy analysis to improve early detection and risk stratification in *BRCA1/2-carrier*s.

**Significance Statement:** Improved clinical surveillance methods are urgently needed for *BRCA1/2* germline carriers. We show that integrating cell-free DNA genomic and epigenomic (fragmentomic and DNA methylation) based assays identifies cancer-associated signals not captured by standard methods, supporting its use as a complementary non-invasive strategy for longitudinal monitoring in high-risk individuals.

## Introduction

Hereditary Breast and Ovarian Cancer (HBOC) syndrome, caused by a pathogenic germline *BRCA1* or *BRCA2* variant, increases the risk of developing various types of cancer, most notably breast and ovarian but also including prostate, pancreatic, and melanoma (1). While the specific risks for each cancer vary depending on *BRCA1* or *BRCA2* carrier status, both female and male carriers approach a lifetime risk of >60%, with the highest risk of breast and ovarian cancer in female *BRCA1* carriers (1–7).

Given the increased cancer risk, individuals with a *BRCA1/2* germline predisposition are advised to undergo high-risk clinical surveillance which include imaging (e.g., mammography and MRI for breast cancer), physical examinations, and prostate specific antigen test for prostate cancer (2,7,8). However, these strategies face several limitations, including limited diagnostic sensitivity, high costs, logistical challenges, and psychological burden in patients (8). There are currently no effective surveillance strategies for ovarian and pancreatic cancers (8). As a preventive strategy, risk-reducing surgeries such as bilateral mastectomy and salpingo-oophorectomy are commonly offered to *BRCA1/2-carriers* (3,9).

Blood-based liquid biopsy offers a minimally invasive approach for cancer surveillance through plasma cell-free DNA (cfDNA) analysis. In cancer, DNA fragments released by tumor cells are referred to as circulating tumor DNA (ctDNA) (10). Tumor-specific alterations including genomic and epigenomic, particularly fragmentomic, analysis of cfDNA have been rapidly advancing for diagnosis, monitoring, tracking treatment response and detecting residual disease (10,11). Historically, analyses of ctDNA have been performed using targeted panel sequencing (TS) to detect variant allele fractions as low as 0.1% (12,13), and shallow (∼1X) whole genome sequencing (sWGS) to detect copy-number alterations at frequencies as low as 3% (14). More recently, sWGS has been employed for fragmentomics, the study of physical and positional characteristics of cfDNA fragments, including sequence motif, length, genomic position and coverage patterns. Fragmentomic approaches can enhance mutation detection (15), predict cancer status (16), and determine the tissue-of-origin from non-genetic chromatin features (17). Many of these concepts have been explored to similar effect in DNA methylation-based cfDNA assays such as cell-free methylated DNA immunoprecipitation and high-throughput sequencing (cfMeDIP-seq) (18), bisulfite-treated (19), or enzymatically-converted cfDNA (20). While ctDNA detection has been broadly evaluated for sporadic cancers, few studies have investigated ctDNA detection in hereditary cancer syndromes (21–23) such as HBOC, with fewer having explored an integrated analysis approach for ctDNA detection.

In this study, we evaluated multimodal cfDNA analysis for early cancer detection in 194 blood samples from 88 *BRCA1/2-*carriers. Using a combination of TS, sWGS and cfMeDIP-seq, we evaluated genomic (somatic variants, copy number alterations), fragmentomic (fragment length, fragment ratio, nucleosome position, and nucleosome accessibility), and epigenomic features independently and as part of an integrated framework to improve early detection. We found that cfDNA could not only confirm clinically diagnosed cancers but also enabled earlier detection than current surveillance strategies in *BRCA1/2-carriers*.

## Results

### Patient cohort

We collected 194 blood samples from 88 unrelated *BRCA1/2-carriers* with confirmed germline testing (age range=31-74 [median=53], germline mutation: *BRCA1*=57, *BRCA2*=28, both *BRCA1* and *BRCA2*=3) from clinics at the Princess Margaret Cancer Centre (Toronto, Ontario) (n=81) and McGill University Health Centre (Montreal, Quebec) (n=7). Male *BRCA1/2-carriers* (patients=2, samples=3) were less represented within the cohort compared to females (n=86, samples=191), likely due to disproportionate participation in surveillance programs (24,25). We classified *BRCA1/2-carrier* samples into four groups: (i) *BRCA1/2-carriers* with active cancer, with no history of a previous cancer [“Cancer Positive (First Positive)” (CP-FP), individuals=25, samples=29], (ii) *BRCA1/2-carriers* with active cancer with a history of previous cancer [“Cancer Positive (Survivor)” (CP-SY), individuals=39, samples=58], (iii) *BRCA1/2-carriers* who have never had cancer [“Cancer Negative (Survivor No)” (CN-SN), individuals=10, samples=19], and (iv) *BRCA1/2-carriers* with no active cancer, but with a history of a previous cancer [“Cancer Negative (Survivor)” (CN-SY), individuals=50, samples=88] (Figure 1A).

**Figure 1:**
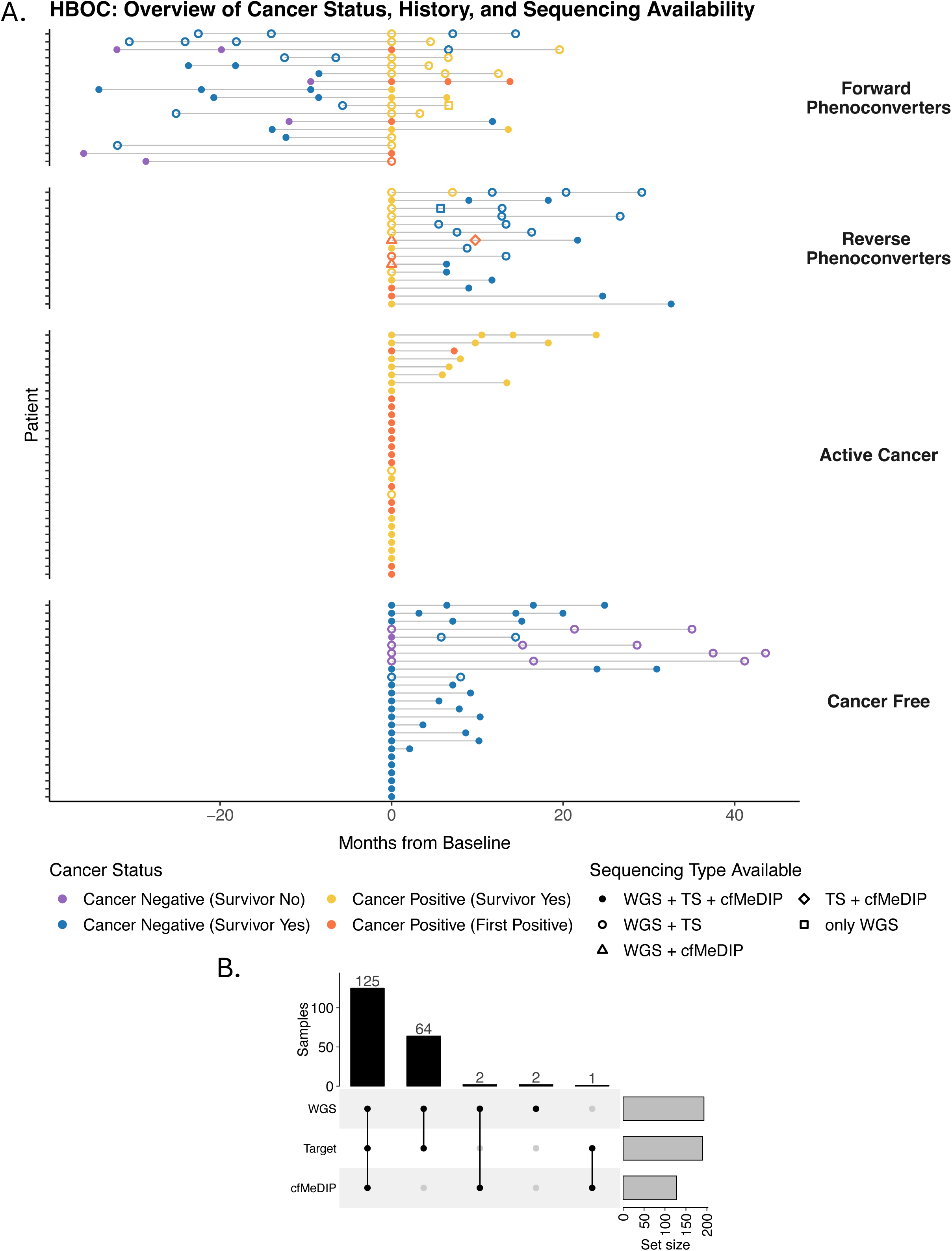
A) Swim plot illustrating patient blood draw timeline. Each horizontal bar represents an individual patients, grouped into Forward Phenoconverters (transitioned from cancer-negative to cancer-positive), Reverse Phenoconverters (cancer-positive to cancer-negative), Active Cancer (only cancer-positive), and Cancer Free (only cancer-negative). Symbols denote the sequencing type available (WGS, TS, cfMeDIP), and are colored by *BRCA1/2-carrier* groups. “Months from Baseline*”* (x-axis) is measured from the first cancer-positive blood draw, or from the first blood draw for patients who were in the Cancer Free group. B) Upset plot showing the number of samples across the different intersections of targeted sequencing (Target) plasma shallow whole genome sequencing (WGS) and Cell-free methylated DNA immunoprecipitation and high-throughput sequencing (cfMeDIP)

Cancer-positive samples were collected from *BRCA1/2-carriers* with a wide spectrum of cancers initially detected through surveillance programs in Ontario and Quebec (mammogram, imaging, and breast exams; see Methods) (2,7). A median of 2 serial blood samples (range=1-5) were collected from 58 *BRCA1/2-carriers*, with the remaining 30 *BRCA1/2-carriers* providing one blood sample (Figure 1A). Among *BRCA1/2-carriers* with serial blood samples, the median time between first and last blood draw was 18.3 months (range=2.1-51.6 months). 32 *BRCA1/2-carriers*, termed “phenoconverters” (Figure 1A), transitioned from cancer-negative to cancer-positive (forward; n=16), cancer-positive to cancer-negative (reverse, n=15), or developed multiple cancers (n=1). A total of 107 cancer-negative (from CN-SN and CN-S) and 87 cancer-positive (from CP-FP and CP-SY) samples were collected. Sample descriptions can be found in Supplemental Table 1. A total of 45 healthy control plasma samples were also collected. Profiling by all three protocols: sWGS, TS, and cfMeDIP-seq was performed for 125 samples, two methods for 64 (sWGS/TS), 2 (sWGS/cfMeDIP), and 1 (TS/cfMeDIP) samples; and one method (sWGS) for two samples (Figure 1B). All 45 samples from healthy controls were profiled by sWGS.

### Verification of germline *BRCA1/2* alterations in cfDNA

We verified pathogenic *BRCA1/2* germline variants in all 190 samples from 88 patients, including all variants from clinical reports available from 82 patients. Medical records from six patients were inaccessible. Sequence variant calling in cfDNA samples with matched normal buffy coat identified *BRCA1/2* germline single nucleotide variants (SNVs) or in/dels in 81 patients (samples=172) (Supplemental Table 2). For cfDNA samples in which a germline sequence variant was not identified (patients = 6), we performed exon-level copy-number analysis which identified a germline deletion in all six patients: full-gene deletion (n=1), multi-exon deletion or duplication (n=2), and a single exon deletion (n=3). Finally, one patient for whom a pathogenic germline alteration was not detected, we performed manual inspection of the raw reads which identified a 3-base deletion (*BRCA1*, c.5062_5064delGTT). This demonstrates that pathogenic *BRCA1/2* variants are readily detected in cfDNA without the need for additional cell-based testing.

### Detection of somatic mutations and genome-wide copy number alterations in cfDNA

To assess the frequency of secondary, somatic alterations in cfDNA, we performed matched cfDNA/matched normal buffy coat calling of analysis of TS data from 190 samples from 88 patients. This identified a somatic variant in 24 samples from 12 patients: 15/84 Cancer Positive timepoints (17.9%) and 9/106 Cancer Negative timepoints (8.5%). Among the 27 CP-FP samples, we identified four mutations across four patients (4/27; 14.8% detection rate): 1 *BRCA2*, 2 *TP53*, and 1 *PALB2* (Figure 2). In the 57 CP-SY samples, we identified 11 mutations across seven patients (11/57: 19.3% detection rate): 2 *BRCA1*, 8 *TP53*, and 1 *PALB2*. Among 19 CN-SN samples, we identified a recurrent *TP53* p.Ser90LeufsTer59 mutation in serial samples from one patient (2/19: 10.5% detection rate). Among CN-SY, we found 7 mutations from 4 patients (7/87; 8.0% detection rate): 3 *BRCA1*, and 4 *TP53*. Interestingly, all four cancer-negative patients with a somatic mutation detected in cfDNA (samples=8) would go on to develop cancer (11, 18, 18, and 32 months later). One additional cancer-negative patient with a somatic mutation in cfDNA was lost to follow-up, so they were not included in further analysis.

**Figure 2:**
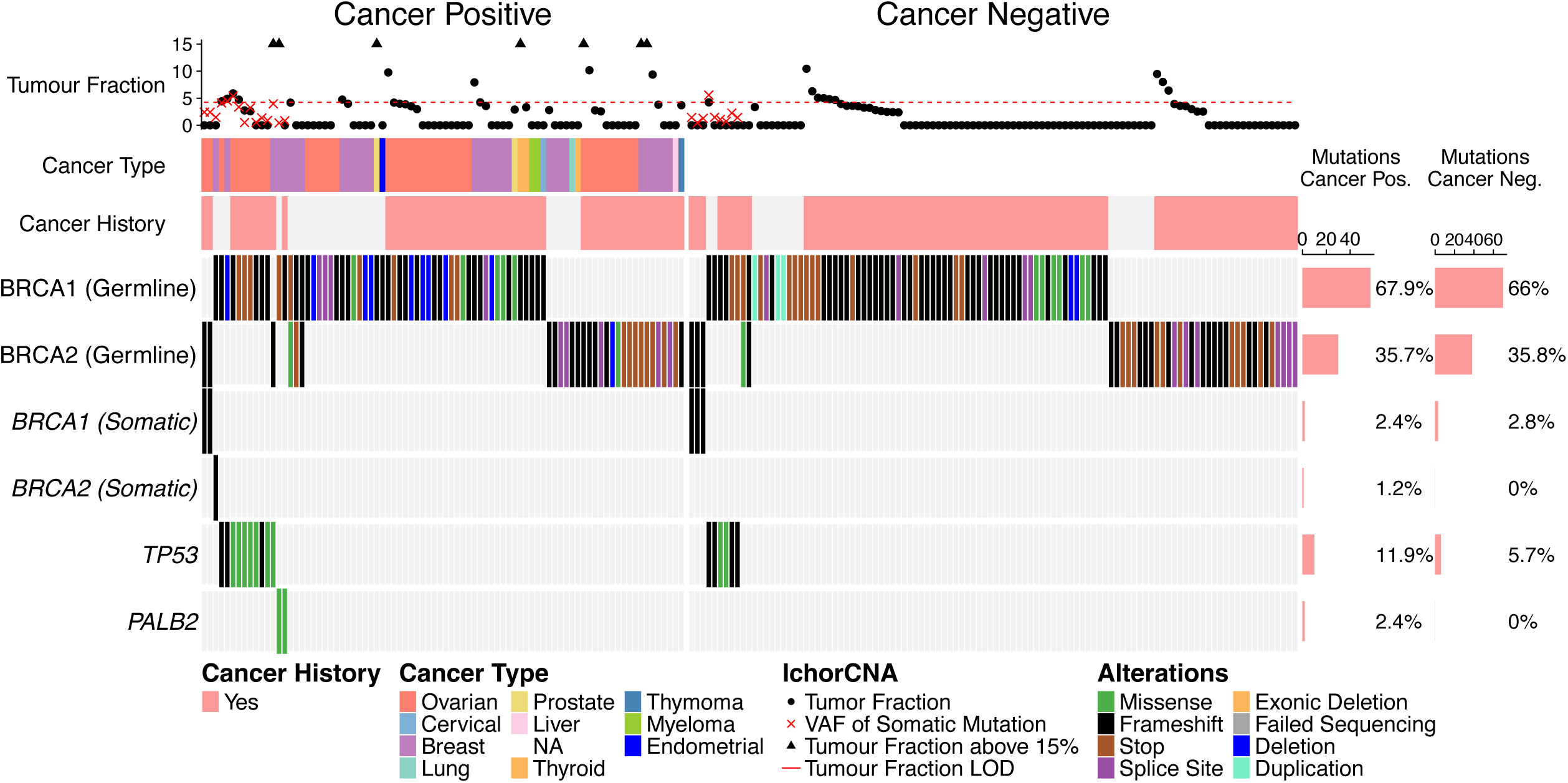
Oncoplot showing germline and somatic variants identified in the plasma of *BRCA1/2-carriers* using targeted panel sequencing. Mutations (rows) include germline and somatic *BRCA1* and *BRCA2*, *TP53* and *PALB2*. Variant allele frequency (VAF) of detected somatic mutation, Copy-number informed tumor fraction profiled using sWGS, and clinical information is displayed at top. Barchart displaying percentage (%) of cancer-positive and cancer-negative *BRCA1/2-carrier* samples with a mutation in each gene are on the right.

To assess the impact of genome-wide detection of copy number alterations (CNAs) in cfDNA, we applied the ichorCNA algorithm (13) to sWGS (downsampled to median 2.1x) from 193 samples (patients=88). We identified 25 samples (patients=21) that had a detectable tumor fraction (TF) of >= 4.2%, a limit of detection (LOD) based on the 99th percentile of 45 healthy controls. We detected CNAs in 19% of CP-SY samples (11/58), 18% of CP-FP samples (5/28), and 10% of CN-SY samples (9/88) (Supplemental Figure 1A; Supplemental Table 3). Of 189 samples (patients=88) with both TS (variant calling) and sWGS (CNA) (CP-FP=26, CP-SY=57, CN-SY=87, CN-SN=19), 43 samples (26 patients) showed a detectable TF or somatic mutation (18 mutations-only, 19 CNVs-only, six had both). Using this combined approach, we detected 25/83 Cancer Positive cases (30%) and found evidence of cancer in 18/106 Cancer Negative cases (17%) (Supplemental Figures 1B,C). Reclassifying cancer-negative samples with a cancer detected within 12 months as cancer-positive, combined somatic mutation and genome-wide CNA detection achieved 28% sensitivity, 83% specificity, 51% negative predictive value (NPV) and 64% positive predictive value (PPV). These data suggested a need for additional molecular features to improve the limit of detection and NPV for cancer surveillance.

### Genome-wide cfDNA fragment length profiles distinguish cancer status in *BRCA1/2-carriers*

Our previous study demonstrated that individuals with Li-Fraumeni syndrome (LFS), exhibit an increased proportion of short cfDNA fragments compared to healthy controls (21), motivating us to assess whether similar patterns occur in *BRCA1/2-carriers*. While all *BRCA1/2-carriers* and Healthy Controls exhibited the expected ∼167 bp peak (median=169bp; Supplemental Figures 2A,B), only *BRCA1/2-carriers* cancer survivors (CP-SY and CN-SY) showed a statistically significant increase in sub-nucleosomal fragments (10-150 bp). Across all *BRCA1/2-carrier* groups, we observed a decreased proportion of mono-nucleosomal fragments (10-150 bp) and increased di- and tri-nucleosomal fragments (300-600 bp) compared to healthy controls (p-value<0.05; Supplemental Figure 3A). These fragmentation profiles did not differ by cancer status (Supplemental Figure 3B; p>0.05), suggesting that these alterations are intrinsic to *BRCA1/2-carrier* status, potentially masking generalized cancer-specific fragment shortening.

We next investigated genome-wide fragmentation differences in *BRCA1/2-carriers* using DELFI comparing relative abundance of short (90–150 bp) to long (151–220 bp) fragments in 5 Mbp bins across the genome (16) (Figure 3A). Our analysis revealed a ∼1.25x increased standard deviation in the *BRCA1/2-carriers* profiles compared to healthy controls, independent of cancer status (CN-SN=1.18 sd, CN-SY=1.20 sd, CP-FP=1.28 sd, CP-SY=1.38 sd) (Supplemental Figure 4).

**Figure 3:**
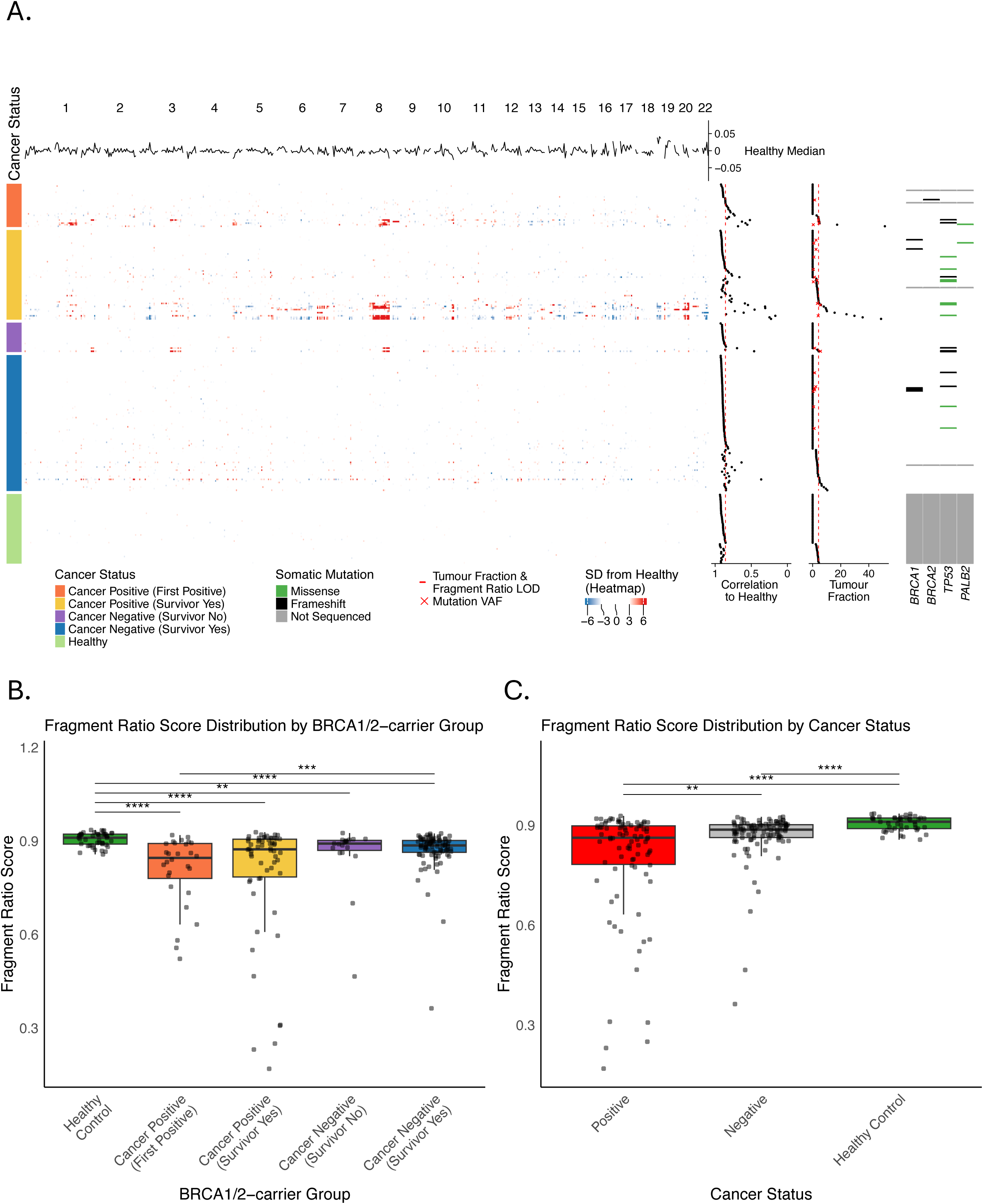
A) Heatmap of genome-wide fragment ratio profiles (90-150 bp/151-220 bp) of healthy controls and *BRCA1/2-carriers* separated by cancer status group are shown on the left. Pearson’s correlation scores (Fragment Ratio Score) of *BRCA1/2-carriers* compared to healthy control median, copy-number informed tumor fraction and somatic mutations (from targeted sequencing) shown on the right. Tukey boxplots comparing Fragment Ratio Score across (B) cancer status groups and healthy control and (C) cancer status and healthy control. P-values were calculated using a two-sided Mann-Whitney U test (Wilcoxon rank-sum test) between all pairwise combinations and adjusted for multiple testing using the Holm method. Only statistically significant comparisons (adjusted p < 0.05) are displayed. * = adj. p < 0.05, ** = adj. p < 0.01, *** = adj. p < 0.001, **** = adj. p < 0.0001.

To assess per-sample fragmentation in *BRCA1/2-carriers*, we defined a Fragment Ratio Score (FRS): the Pearson correlation between each sample’s profile and the median profile of the 45 healthy controls (left box plot Supplemental Figure 4). Consistent with shorter fragments corresponding to large somatic copy number alterations, cancer-positive samples exhibited the lowest score (CP-FP median 0.85, range 0.52-0.92; CP-SY median 0.87, range 0.17-0.93). Cancer negative scores correlated more strongly with healthy controls (CN-SY median 0.89, range 0.36-0.92; CN-SN median 0.89, range 0.47-0.93), none exceeding 0.93. This supports past work suggesting intrinsic fragmentomic differences between hereditary cancer carriers and non-carriers.

Comparing within *BRCA1/2*-carriers, FRS were significantly higher in the cancer-positive versus the cancer-negative group, independent of prior diagnosis (Wilcoxon rank-sum p-value<0.003; Figure 3B,C). Fragment Ratio Scoring identified 63 molecular-positive samples (patients=46), resulting in detection rates of 61% (17/28) in CP-FP, 41% (24/58) in CP-SY, 23% (20/88) in CN-SY, and 16% (3/19) in CN-SN. The observed fragmentation differences may reflect underlying changes in chromatin architecture and transcriptional activity in cancer cells, supporting their potential as genomic and epigenomic markers for early cancer detection (16,26).

### Increased intra-nucleosome fragmentation in cancer survivors without an active cancer

The characteristic non-random fragmentation patterns of cfDNA, resulting from nucleosome protection during DNA degradation, led us to investigate whether we could infer the origin of cfDNA fragments in BRCA1/2-carriers cfDNA. To first assess fragmentation patterns against normal blood cells that would contribute the greatest cfDNA signal, we mapped cfDNA fragment start and end points from all BRCA1/2-carriers and healthy controls relative to a reference set of ∼13 million nucleosomes derived from peripheral blood cells (27). As anticipated, all samples showed an M-shaped fragment-end distribution with peaks at +/-83 bp from the nucleosome centers (∼167 bp fragment length) (27,28). However, BRCA1/2-carrier groups exhibited decreased fragment-end frequency at the peaks and increased frequency within the nucleosome spanning regions compared to healthy controls (Kolmogorov–Smirnov p-values: < 0.0000011–0.0001; Supplemental Figure 5A), consistent with our experience in the LFS hereditary cancer syndrome (22). These observations suggest a significant contribution of cfDNA fragments inherent to *BRCA1/2-carriers* that do not correspond to the non-carrier PBMC reference.

To assess whether these differences reflected active cancer in *BRCA1/2-carriers*, we calculated Nucleosome Peak Scores (NPS) by summing z-scores (vs. control median) across the ±83 bp nucleosome region (See Methods). No significant differences were observed between cancer-positive and cancer-negative carriers (Wilcoxon rank-sum p>0.49; Supplemental Figure 5C). However, scores were significantly elevated in cancer-positive and survivor samples (CP-FP, CP-SY, CN-SY) compared to healthy controls (p-value<0.0005-0.023), while cancer-negative carriers without a prior cancer (CN-SN) did not differ (p-value>0.54) (Supplemental Figure 5B). NPS identified 49 molecular-positive samples (patients=33), with detection rates of 29% (8/28) in CP-FP, 28% (16/58) in CP-SY, 28% (25/88) in CN-SY, and 0% (0/19) in CN-SN. This analysis opens the possibility of lifetime risk prediction for developing cancer from intrinsic fragmentation patterns and tailoring of cancer surveillance frequency using cfDNA.

### Altered chromatin accessibility in active cancer *BRCA1/2-carriers*

Given the observed genome-wide dysregulation of fragments, we hypothesized that nucleosome accessibility, inferred from fragmentation patterns, could offer insights into the specific cancer subtypes encountered in our heterogeneous cohort. To test this hypothesis we employed Griffin (17), a computational framework to differentiate cancer states via chromatin accessibility. Analyzing ATAC-seq peaks derived from 23 TCGA cancer types and an in-house ovarian cancer organoid dataset (see Supplemental Methods), we assessed the Griffin midpoint coverage and amplitude of each sample for each cancer ATAC-seq reference. Our primary objective was to assess if cancer-positive *BRCA1/2-carriers*, particularly breast and ovarian cancers, exhibited increased chromatin accessibility (decreased midpoint coverage) consistent with reference profiles. Only a subset of cancer-positive samples, including breast and ovarian, closely aligned with their matched cancer type, characterized by decreased midpoint coverage and higher amplitude. Interestingly, several cancer-negative *BRCA1/2-carrier* samples also displayed increased accessibility patterns aligning with specific cancer types (Figure 4A, Supplemental Figure 6A). Since we did not observe distinct cancer-type-specific midpoint coverage patterns, we computed a midpoint coverage z-score for each sample across the 24 cancer types (see Methods; Supplemental Figure 6B). This analysis revealed that the reference-based ATAC-seq approach preferentially detected samples with strong cancer-associated chromatin signals, but exhibited low overall sensitivity. Positive z-scores across multiple ATAC-seq references suggest broad rather than cancer-type-specific signal detection, potentially confounded by *BRCA1/2*-associated fragmentation patterns.

**Figure 4:**
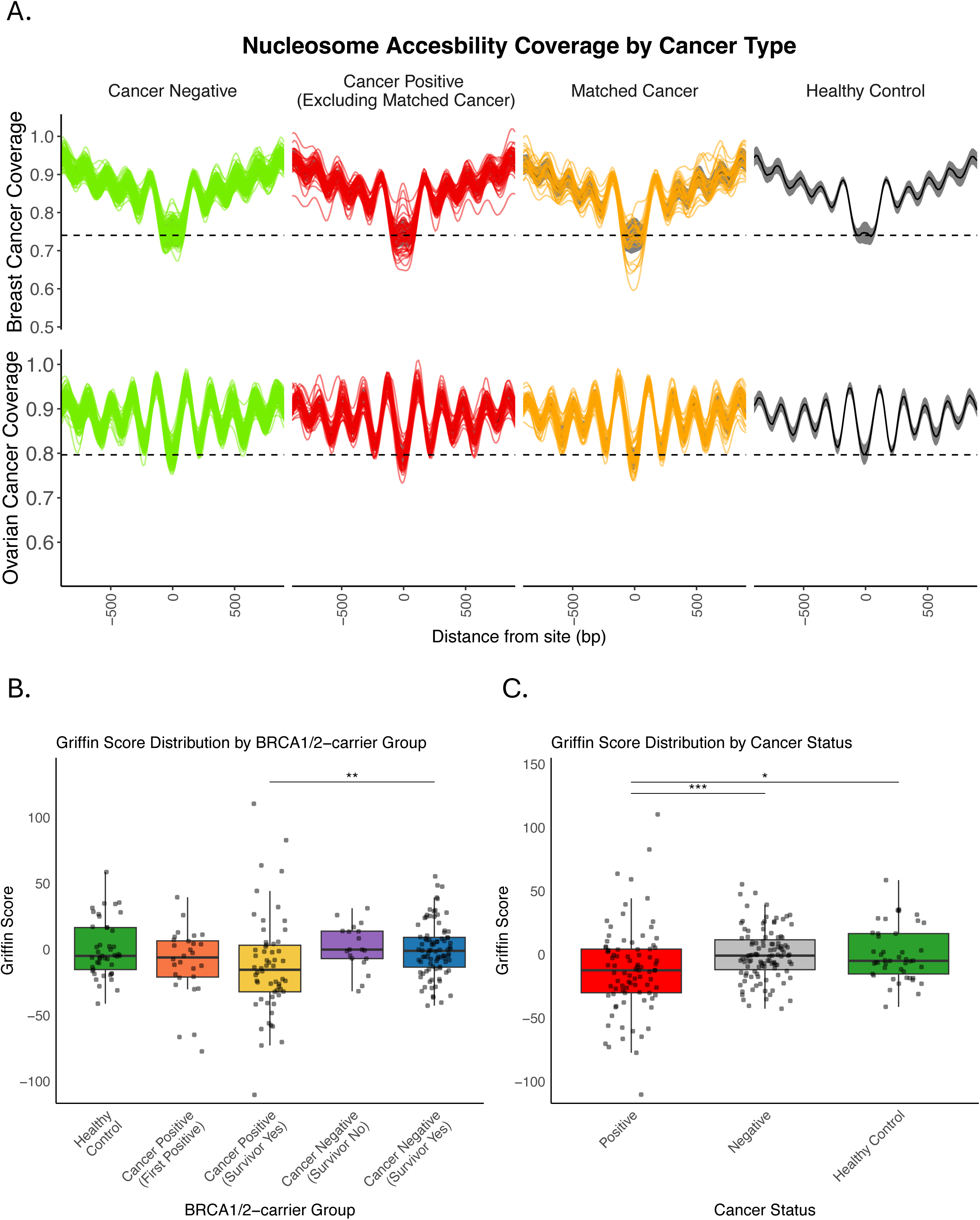
A) Nucleosome Accessibility tracks showing chromatin accessibility at breast cancer (top) and ovarian cancer (bottom) associated open chromatin sites. Samples from Cancer Negative, Cancer Positive (Excluding Matched Cancer), Cancer Positive (Matched Cancer) are displayed in each respective plot. The respective cohort medians are displayed in black +/− 1 standard deviation. Tukey boxplots comparing Consensus Griffin Score across (B) cancer status groups and healthy control and (C) cancer status and healthy control. P-values were calculated using a two-sided Mann-Whitney U test (Wilcoxon rank-sum test) between all pairwise combinations and adjusted for multiple testing using the Holm method. Only statistically significant comparisons (adjusted p < 0.05) are displayed. * = adj. p < 0.05, ** = adj. p < 0.01, *** = adj. p < 0.001, **** = adj. p < 0.0001.

Due to the non-specific midpoint coverage across cancer types, we developed a Consensus Griffin Score (CGS) by summing the midpoint z-scores across the 24 cancer-type reference profiles (see Methods). CGS identified 17 molecular-positive samples (patients=14), resulting in detection rates of 11% (3/28) in CP-FP, 21% (12/58) in CP-SY, 2% (2/88) in CN-SY, and 0% (0/19) in CN-SN. Cancer-positive samples exhibited a general increase in CGS, rather than specific tumor subtyping via nucleosome accessibility (Figure 4B,C). These findings suggest that nucleosome accessibility shifts in *BRCA1/2-carriers* are cancer-history dependent, and become more dysregulated with an active cancer.

### Uni-modal scoring analysis using static thresholds for cfDNA metrics

Our genomic and epigenomic (fragmentomic) approaches detected cancer-associated signals in *BRCA1/2-carriers*. While some methods distinguished cancer-positive from cancer-negative samples and, in some cases, reflected prior cancer history, no singular approach was perfect in all cases. We therefore tested whether models integrating genomic and epigenomic features from the fragmentomics data could improve sensitivity for detecting cancer-associated signals.

Among 83 cancer-positive samples (patients=61) profiled by both TS and sWGS, at least one ctDNA signal was detected in 55 samples (66%) (patients=43): 3 genomic-only (mutation or CNV) only, 30 fragmentomic only, and 22 showing both. Detection rates per feature included somatic variants (15/83; 18%), copy number variations (CNVs, 16/83; 19%), short fragment enrichment (FS, 23/83; 28%), fragment ratio shifts (FRS, 40/83; 48%), decreased midpoint coverage (CGS, 40/83; 48%), and altered nucleosome peak positioning (NPS, 23/83; 28%) (Figure 5, Supplemental Figure 7). Notably, detection rates were similar across cancer types: 26/39 (67%) for ovarian and 23/31 (74%) for breast cancers.

**Figure 5:**
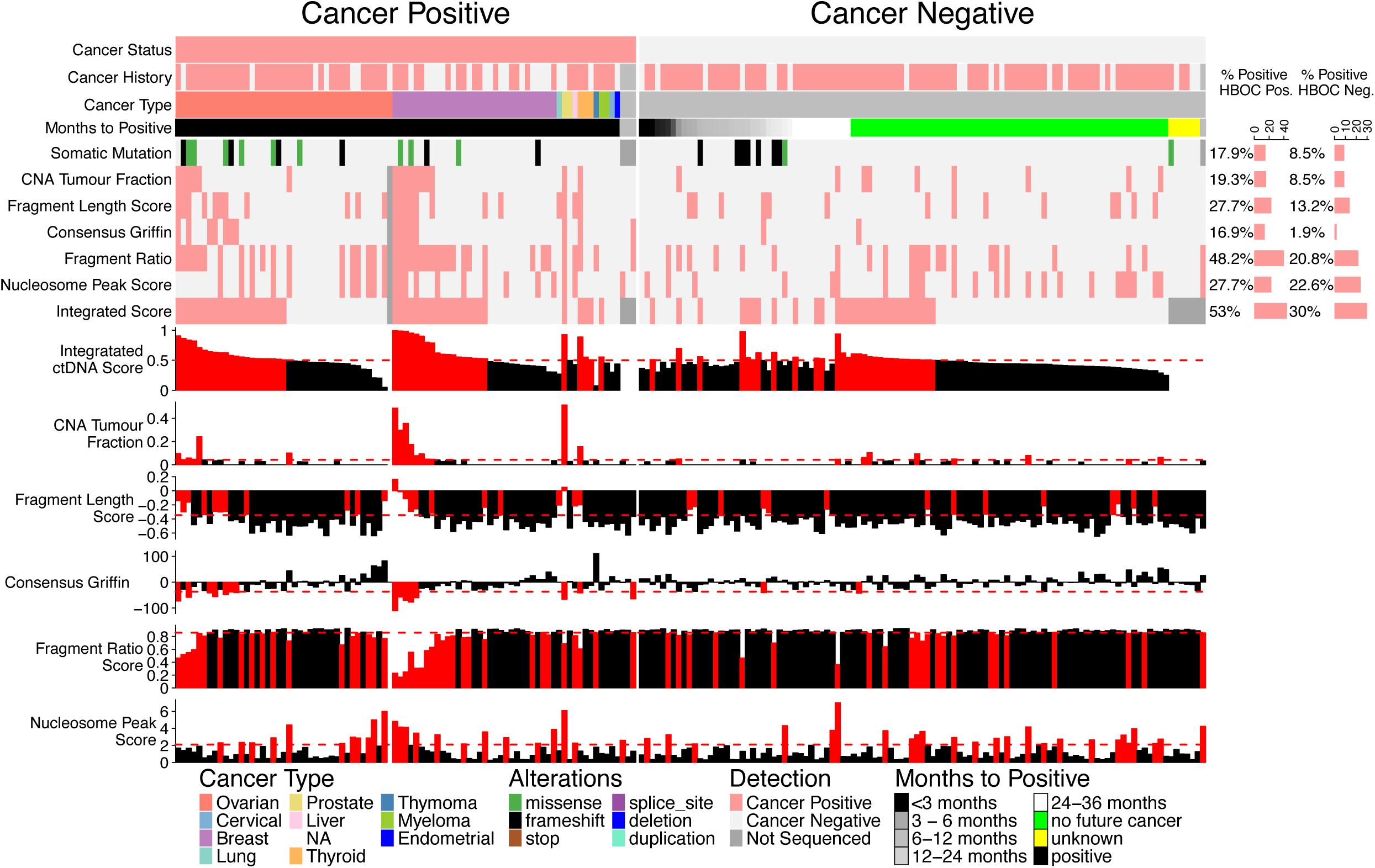
Heatmap and barcharts displaying each genomic, fragmentomic and integration scores. Additional clinical information is displayed at the top (cancer status, cancer history, cancer type, months to next positive timepoint). Barchart displaying percentage (%) of cancer-positive and cancer-negative *BRCA1/2-carrier* samples with a molecular positive status for each score are on the right.

ctDNA signals were also detected in cancer-negative samples, suggesting either false positives or clinically undetected cancer. Among 100 cancer-negative samples (patients=54), 49 samples (49%) (patients=30) were ctDNA positive: somatic variants (8/100; 8%), CNVs (9/100; 9%), fragment ratio (22/100; 22%), nucleosome peak (24/100; 24%), fragment length (14/100; 14%), and Griffin score (2/100; 2%). Clinical follow-up (“Time to Next Positive”; Figure 5) revealed that 13 of the 30 (43%) ctDNA-positive cancer-negative patients (43%; 20 samples) developed cancer, including 6 samples (5 patients) within 12 months. Notably, 12 patients belonged to the CN-SY group, suggesting that ctDNA positivity may reflect residual disease or early recurrence. This suggests that the false-positive rate is 29% (29/100) within samples and 32% (17/54) within patients, highlighting the need for clinical verification of any positive cfDNA signals in practice.

To assess negative ctDNA time points and patients, we considered a true negative as validated if the patient did not develop cancer within 12 months of the negative ctDNA result. Using this metric, within cancer-negative *BRCA1/2-carrier* samples, unimodal PPV values range from 22% to 100% and NPV ranged from 86% to 88% (Supplemental Figure 8). With detection by any method (>=1 ctDNA signal), PPV was 41% (20/49) and NPV was 86% (40/51). Despite strong NPV performance appropriate for surveillance, the relatively low PPV emphasizes the need for confirmatory imaging to guide clinical follow-up.

### Multimodal approach to combine continuous cfDNA metrics

Building on the unimodal, static-threshold analysis, we next examined whether integrating multiple cfDNA features could improve detection accuracy. Notably, 35 of 83 cancer-positive samples (42%; 28 of 61 patients) and 25 of 100 cancer-negative samples (25%; 17 of 54 patients) exhibited at least two ctDNA signals. This suggests that the co-occurrence of multiple abnormal signals may be more indicative of cancer status than any single-feature threshold.

To test this hypothesis, rather than applying static thresholds that impose rigid constraints on feature integration, we employed a machine learning strategy. We trained four distinct algorithms including k-nearest neighbors (KNN), generalized linear models (GLM), support vector machines (SVM), and random forest (RF) on our six continuous cfDNA features: somatic mutation VAF, TF measured by ichorCNA, NPS, CGS, FS, and FRS. When comparing healthy controls and cancer-negative *BRCA1/2-carriers*, models achieved high classification performance with mean AUC-ROCs ranging from 0.765 to 0.842 (95% CI: 0.728–0.866) on the test set (Supplemental Figure 9A). Model performance in distinguishing 83 cancer-positive individuals from a subset of 30/60 cancer-negative individuals who never developed cancer was modest, with mean AUC-ROCs ranging from 0.600 to 0.649 (95% CI: 0.566–0.688) on the test set (Supplemental Figure 9B). Although specificity was reduced, the models achieved NPVs ranging from 0.759 to 0.835 (95% CI: 0.718–0.875), suggesting that multi-feature integration may enhance sensitivity. To evaluate whether these models could aid in early detection of the remaining 30/60 cancer-negative samples, we selected the best-performing pre-trained models. Using the optimal thresholds from the training set, determined by the Youden index, we applied them to the held-out validation set comprising 40 cancer-negative individuals who later developed cancer (true positives) and 30 who did not (true negatives). Although overall performance declined (AUC-ROC range: 0.498 – 0.622), the RF model achieved the highest AUC-ROC value (0.622; PPV=61%, NPV=52%). The PPV indicates that 61% of samples predicted to develop cancer were correctly identified. However, the low NPV of 52% reveals limited reliability in ruling out future cancer risk within this cohort, underscoring the complexity of distinguishing *BRCA1/2-carriers* samples across heterogeneous disease states and clinical progression timelines.

### Sample-level scoring based on integration of cfDNA features

To complement the machine learning models and provide a more clinically interpretable sample-level metric, we developed a *BRCA1/2*-specific integration score for cancer status (positive vs. negative) using logistic regression (see Methods), following our previous approach in our LFS cohort (21). This score incorporated all six cfDNA features: somatic mutation VAF, TF, FS, FRS, CGS, and NPS. It was applied to *BRCA1/2-carrier* samples with complete TS and sWGS data and known clinical outcomes, excluding those with unknown time to next cancer diagnosis (samples=183). A molecularly positive score (LOD>0.5) was observed in 44/83 (53%) cancer-positive and 30/100 (30%) cancer-negative samples. Importantly, 14 of the cancer-negative samples developed cancer within 5.2 years (median 2.2 years), including 3 within 12 months. Integration scores were significantly higher in the cancer-positive compared to the cancer-negative cohort (p-value<0.00036; Figure Supplemental 10B), with the strongest signals observed in the CP-SY samples compared than CN-SY (p-adjusted<0.0037) (Figure Supplemental 10A). Together, these results suggest that the integration score captures meaningful biological signals and translates them into a complementary, clinically interpretable measure that can be applied to individual samples for cancer risk assessment in *BRCA1/2-carriers*.

### Longitudinal sampling for early detection

To explore longitudinal ctDNA trends, we assessed the clinical imaging and 7 cfDNA features from 3-5 samples collected over 18-52 months for 4 select patients (Figure 6).

**Figure 6:**
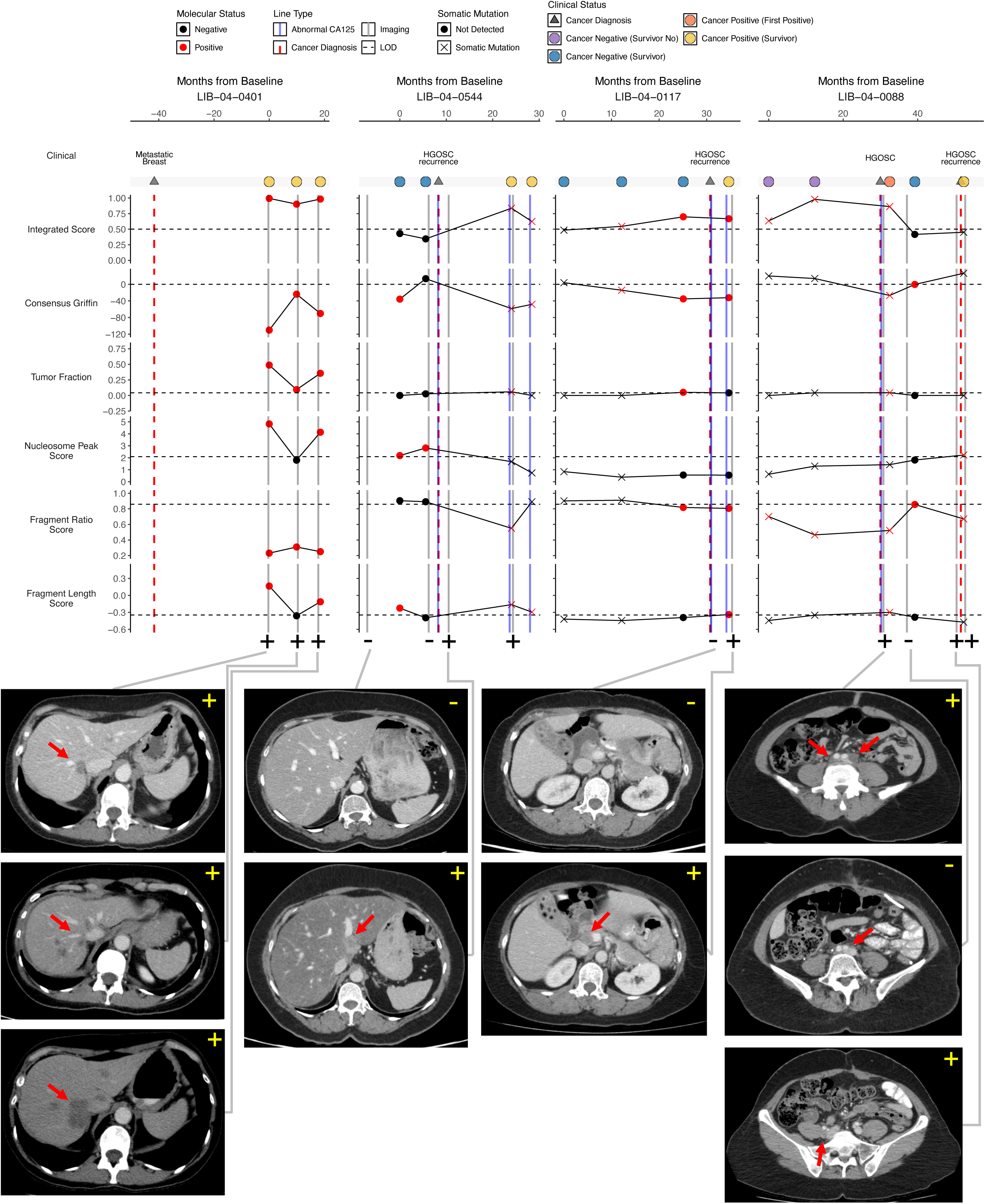
Longitudinal tracks for select *BRCA1* or *BRCA2* patients showing ctDNA signal, clinical information, and imaging data.

Exemplar “always positive” patient: Patient **LIB-04-0401** had metastatic breast cancer over a 60-month period during which imaging remained positive and three blood draws were collected. We detected elevated ctDNA signals across all draws (6/7 molecular positive; no detectable mutation). Notably, the second blood draw showed no significant nucleosome peak score or fragment length score (FS) coinciding with imaging that indicated improvement in metastatic disease (4/7 molecular positive). Conversely, at the third timepoint, disease progression was evident in imaging that was mirrored by increased ctDNA signal levels (6/7 molecular positive). Overall, ctDNA findings were consistent with clinical imaging and indicative of persistent, active cancer.

Patient **LIB-04-0544** had 4 blood draws over 28 months. A abdomen/pelvis CT scan 15 months before HGSOC recurrence was negative. At 8 months pre-recurrence, we detected significant signals in 3/7 cfDNA features, including 3 fragmentomic scores. Three months before diagnosis, only the nucleosome-peak metric remained positive, with the patient having a negative abdominal ultrasound 2 months prior to diagnosis. Subsequent increases in CA-125 levels and abdominal symptoms prompted thoracic and abdominal CT imaging, which confirmed recurrence. At 15 months post-recurrence, abdomen/pelvis CT imaging revealed progressive disease, with 6/7 ctDNA features testing positive, including the emergence of a novel somatic *TP53* mutation (c.379T>A), consistent with concurrent elevations in CA-125.

Patient **LIB-04-0117** had four blood draws over a 34-month period. We detected a somatic *TP53* mutation (c.985del) 31 months prior to HGSOC diagnosis, while all other ctDNA metrics remained negative. We then detected a positive integration and Consensus Griffin Score 19 months prior to diagnosis. Six months before diagnosis, additional cfDNA features, including copy-number-informed tumor fraction and FRS, also became molecular positive. At time of diagnosis (30 months after the baseline blood draw), the patient presented with abnormal CA125 levels, but abdominal CT imaging was unremarkable. The diagnosis was ultimately confirmed 4 months later by thoracic and abdomen/pelvic imaging, supported by ctDNA-based detection through a positive integration score and three additional fragmentomic features.

Patient **LIB-04-0088** had five blood draws over a 51-month period. An abnormal integration score was observed at 30 and 18 months prior to HGSOC diagnosis, likely driven by elevated fragment ratio, and detection of a somatic *TP53* mutation (c.263_264insG). The integration score and somatic mutation remained positive at diagnosis, accompanied by 3 additional fragmentomic signals (6/7 ctDNA positive features). At 8.5 months post-diagnosis, when the patient was clinically cancer-free, only 2/7 ctDNA positive features remained and the somatic mutation was no longer detectable. 14 months later, at the subsequent and final blood draw, CT imaging of the thorax, abdomen, and pelvis confirmed HGSOC recurrence, with 3 out of 7 ctDNA features testing positive and the re-detection of the *TP53* somatic mutation.

### Prediction of cancer of origin using follow-up DNA methylation profiling

While ctDNA integration scores enabled detection of cancer-related signals in *BRCA1/2-carriers*, they lacked specificity for identifying the tissue of origin. To improve tumor-type resolution, we performed cfMeDIP-seq on integration score-positive samples. Due to limited representation of other tumor types in our retrospective cohort, only breast and ovarian cancers were evaluated.

We built a breast cancer–specific DNA methylation classifier by training a random forest model using the differentially methylated regions (DMRs) in plasma samples from active breast cancer (n=25) against samples from cancer-negative (n=69), non-breast cancer (n=40), and healthy controls (n=30). The model achieved an average AUC of 0.786 (95% CI: 0.78–0.793) across 100 rounds of cross-validations in classifying breast cancers from other non-breast cancer samples (Supplemental Figure 11A). Feature importance analysis further identified a signature of 181 DMRs (Materials and Methods, Supplemental Table 4). The same analysis was applied to ovarian cancer (n=26), yielding an average AUC of 0.694 (95% CI: 0.686–0.702) in distinguishing ovarian cancer from other cancer types (Supplemental Figure 11B). 186 DMRs were identified in down-stream analysis as ovarian cancer signatures (Supplemental Table 5).

To predict the tumor of origin for each sample, the average bin-wise probability across breast-specific and ovarian-specific DMRs was calculated to generate breast-positive and ovarian-positive scores, respectively. The 99th percentile of scores from healthy control samples was used as the detection threshold. Among 50 integration-positive samples (Figure Supplemental 11C), the breast score achieved a PPV of 71% and NPV of 89% (10 TP, 4 FN, 4 FP, 32 TN). True positives in this context indicate that the cfMeDIP-seq signal correctly matched the clinically confirmed breast tumor type, supporting the feasibility of inferring the tissue-of-origin for a detected ctDNA signal. The ovarian score yielded a PPV of 35% and NPV of 87% (7 TP, 4 FN, 13 FP, 26 TN). The high false-positive rate suggests that our assay often overcalled ovarian cancer, reflecting limited ovarian specificity. However, clinical follow-up revealed that all nine cancer-negative samples (patients=5) with a positive ovarian score were from previously treated ovarian cancer cases, including three patients who later developed clinically confirmed recurrence. These findings imply that the cfMeDIP-seq signal may capture residual or subclinical disease, highlighting its potential clinical utility for identifying tumor of origin enabling earlier, more precise clinical decision-making.

## Discussion

We investigated the utility of plasma-based liquid biopsy for early cancer detection in patients with HBOC syndrome by retrospectively analyzing 194 blood samples from 88 *BRCA1/2* germline mutation carriers. Our integrated genomic and epigenomic profiling approach demonstrates that multimodal cfDNA analysis can enhance current surveillance methods and support the development of new early detection strategies for cancers lacking effective screening tools.

Detection of somatic mutations in cfDNA is a key strategy in early, non-invasive cancer detection. In our study, we detected somatic secondary mutations in samples from individuals who were clinically cancer-free at the time of blood draw. Clinical follow-up revealed that all such cases subsequently developed cancer with a median time to diagnosis of 1.75 years, underscoring the potential of liquid biopsy to identify early genomic alteration changes that precede clinical diagnosis. However, somatic mutation and CNA analysis alone showed limited sensitivity in detecting clinically confirmed cancers (25/83 samples; 30%), consistent with prior studies in hereditary cancer syndromes that also reported suboptimal detection performance from genomic alterations alone (21).

To address the limitations of cfDNA somatic mutation and copy number profiling, we examined epigenomic features from fragmentomic data as distinct biomarkers. Fragment length analysis identified intrinsic fragment-length alterations in *BRCA1/2-carriers* relative to Healthy Controls; a phenomenon previously encountered in our previous study in LFS (21). Region-specific fragmentation patterns were further captured using our FRS, improving sensitivity and reflecting copy number variants detected in higher tumor burden patients. Additionally, nucleosome peak analysis revealed greater variation in fragmentation patterns in individuals with a prior cancer history, potentially reflecting lasting chromatin remodeling or residual epigenetic alterations from past malignancy. This persistent signal in the absence of active disease presents a challenge in defining true-negative baselines for early detection in high-risk populations. In line with these findings, the Griffin score exhibited the highest specificity (99%) among epigenomic features from fragmentomic data for detecting ctDNA signals in cancer-negative samples with future cancer. This high specificity suggests the ATAC-seq reference based approach effectively captures genuine cancer-associated chromatin accessibility changes. However, its limited performance in pinpointing a single tumor tissue of origin may reflect the distinct biology of *BRCA1/2-carriers* as a high-risk population, whose baseline fragmentation patterns differ from those of sporadic cancers. These findings highlight the need to incorporate *BRCA1/2-carrier*-specific reference data that could improve tissue-of-origin prediction in this high-risk population. A goal of cfDNA-based early detection in HBOC is to identify cancer signals missed by current surveillance methods focused primarily on breast and prostate cancer. Focusing on samples with negative clinical surveillance results, the threshold-based approach for the detection by any method (>=1 ctDNA signal) demonstrated a modest positive predictive value (PPV; 41%) and relatively high negative predictive value (NPV; 86%). For comparison, our previous study of LFS patients (21) using a similar multimodal approach achieved an NPV of 95% and a PPV of 54%. While the NPV observed in this study remains favourable, they are exploratory and must be interpreted with caution. Nonetheless, it is worth noting that current surveillance standards also face limitations. For example, the Ontario Breast Screening Program (OBSP) from 2023 reports a PPV of only 6.2% for MRI and mammography in high-risk populations (29). These findings suggest that despite its modest PPV, cfDNA testing may offer clinical value through frequent, minimally invasive surveillance, with positive results being validated by current gold-standard medical exams and imaging.

While our retrospective study enabled the integration of diverse cfDNA features, it also imposed limitations in terms of sample timing, standardization, and longitudinal follow-up. To advance this work toward clinical implementation, prospective studies are critical. A recent prospective study using fragmentomics for early detection of multiple cancer types demonstrated the potential of this approach, achieving a sensitivity of 53.5% and a specificity of 98.1% in a cohort of 3,724 asymptomatic individuals, albeit with relatively low cancer incidence (43 cases) (30). As our experience showed, longitudinal sampling provides temporal context that can clarify potential false positives. As exemplified by patient LIB-04-0117, their two somatic mutation-positive samples collected 18 and 30 months before diagnosis were classified as false positive, whereas their blood draw 6 months before diagnosis showed a true positive signal based on copy-number informed tumor fraction and FRS, despite the absence of a detectable mutation. Longitudinal sampling may ultimately enable the development of personalized, dynamic machine learning models capable of tracking patient-specific ctDNA trends over time. Such models may offer improved sensitivity by capturing individualized baselines and detecting subtle shifts indicative of early tumor evolution.

Overall, our findings encourage prospective study of multiomic cfDNA profiling for HBOC surveillance. By combining genomic and epigenomic-based signals from fragmentomic and DNA methylation data, this approach enables surveillance for currently-unscreenable cancers, improves early detection sensitivity, and provides a foundation for developing more comprehensive, personalized cancer screening strategies for high-risk populations.

## Materials and Methods

### Study Design and Patient Cohort

This study was approved by the institutional review boards of the University Health Network (UHN) (REB#: 18-5692), and the Jewish General Hospital (JGH) (REB#: MP-05-2020-1928) and conducted in accordance with established ethical guidelines, including the Declaration of Helsinki, CIOMS, the Belmont Report, and the U.S. Common Rule. Written informed consent was obtained from all participants prior to enrolment. All clinical evaluations and treatments were provided by board-certified clinicians according to standard-of-care protocols (see *Supplemental Materials and Methods* for additional details).

### Blood Processing and Extraction

Blood Processing and Extraction were performed as described by Wong *et al.* (21) Detailed protocols are available at https://charmconsortium.ca/protocols-database/. Additional details are provided in the *Supplemental Table 6* and *Supplemental Materials and Methods*.

### DNA Library Preparation and Sequencing for TS and WGS

cfDNA libraries were prepared using the KAPA Hyper Prep Kit (Kapa Biosystems) and xGen Duplex Seq Adapter-Tech Access (IDT). UMI-ligated libraries were split for whole-genome sequencing (WGS) and targeted sequencing (TS).

TS used two hybrid capture panels: CHARM (*TP53, BRCA1/2, PALB2, MLH1, MSH2, MSH6, PMS2, EPCAM, APC*) and REVOLVE (CHARM genes plus *CCNE1, CDH1, VHL, NF1*) (Supplemental Table 7). Dual-indexed libraries underwent hybrid capture and PCR amplification. All libraries were sequenced on the Illumina NovaSeq 6000 (150-bp paired-end), targeting 30X depth for WGS and 20,000X for TS.

Sequencing data were processed as described in Wong *et al.* (21), following established protocols (https://github.com/oicr-gsi/bwa). Briefly, reads were aligned to GRCh38 using BWA (v0.7.12; RRID:SCR_010910), followed by UMI-based deduplication using Samtools (v1.9; RRID:SCR_002105). WGS BAMs were downsampled to 50 million reads (∼1X coverage) using Picard DownsampleSam (v2.10.9). Coverage metrics are provided in *Supplemental Table 8*. Copy Number Alteration and fragmentomic analyses were performed using the PughLab pipeline-suite (https://github.com/pughlab/pipeline-suite).

### Germline and Somatic Mutation Calling in Targeted Sequencing

Targeted sequencing (TS) reads were error-corrected using ConsensusCruncher (https://github.com/pughlab/ConsensusCruncher; RRID:SCR_023654), generating single-strand, duplex, and all unique consensus sequences. Germline variants were called using GATK HaplotypeCaller (v3.8; RRID:SCR_001876) (31) on paired plasma and buffy coat BAMs, restricted to all unique consensus sequences and limited to CHARM panel intervals. Germline CNAs were assessed with PanelCNMops (v1.14.0) (32).

Somatic SNVs were called using Mutect2 (v3.8, Broad Institute; RRID:SCR_026692) (33) on all unique consensus sequences in *BRCA1, BRCA2, PALB2, TP53, MLH1, MSH2, MSH6, PMS2, EPCAM,* and *APC*. Variants classified as germline or shared between plasma and buffy coat were excluded. Additional filters removed benign variants, recurrent artefacts (n>2), and somatic mutations below variant allele frequency (VAF) thresholds (missense/stop <0.1%, frameshift <5%) unless manually reviewed. All retained variants were manually validated in IGV (v2.16.0; RRID:SCR_011793) using matched plasma and normal BAMs. See *Supplemental Materials and Methods* on edge case handling.

### Copy Number Alteration and Tumor Fraction Estimation

Generation of an in-house panel of normals (PON; n=45), copy number alteration (CNA) analysis and tumor fraction estimation were performed using ichorCNA (version 0.2.0; Broad Institute; https://github.com/broadinstitute/ichorCNA) as described by Addalsteinsson *et al.* (13). All analyses, including PON generation and sample processing were executed using the PughLab pipeline-suite (https://github.com/pughlab/pipeline-suite). LOD was set to the 99th percentile of TF in healthy controls (0.0424); samples exceeding this threshold were classified as molecularly positive.

### Fragment Length Analysis and Score Calculation

Global fragment length distributions were computed using Picard CollectInsertSizeMetrics (v4.0.1.2; RRID:SCR_006525), retaining fragment lengths between 1 to 599 bp. Fragment length scores were calculated using a previously published reference set from Vessies *et al.* and described further in Wong *et al.* (21,34). A brief overview of the scoring approach is provided in *Supplemental Methods and Materials*. The LOD was defined as the 99th percentile of healthy control scores (LOD=-0.346); samples exceeding this threshold were classified as molecularly positive.

### Fragment Ratio Analysis and Score Calculation

Fragment ratio was based on the DELFI approach of Cristiano *et al.* (16) and adapted for hg38 reference genome as described by Wong *et al.* (22). Fragment ratio scores were calculated as the Pearson correlation between each sample’s bin-wise profile and the healthy control median. The LOD was set to the 1st percentile of healthy control correlations (LOD=0.859); samples below this threshold were classified as molecularly positive.

### Nucleosome Positioning Analysis and Score Calculation

Nucleosome Positions were analyzed as previously described by Wong *et al.* (22) (see *Supplemental Methods and Materials*). Nucleosome Peak Scores were derived by analyzing cfDNA fragments spanning the nucleosome region (-/+83 bp; ∼167 bp). Bin-wise z-scores were calculated based on the mean of healthy controls and averaged to obtain a final score per sample. The LOD was set as the 99th percentile of scores from healthy controls (LOD=2.1); samples above this threshold were classified as molecularly positive.

### Nucleosome Accessibility and Score Calculation

Nucleosome positions were analyzed using Griffin tool (v0.1.0; https://github.com/adoebley/Griffin) as described in Doebley *et al.* (17) and adapted as previously described by Wong and colleagues (22). We used 10,000 tissue-specific open-chromatin sites from TCGA for 23 cancer types and generated a custom ovarian reference using the top 10,000 peaks from ovarian organoid ATAC-seq data (see *Supplemental Materials and Methods*).

To compute the Consensus Griffin Score (CGS), we calculated z-scores of fragment coverage at five midpoint positions (−30, −15, 0, 15, 30) for each cancer type, relative to the healthy control median. All z-scores across the 24 cancer types were then summed to yield a single CGS per sample. The limit of detection (LOD) was defined as the 99th percentile of CGS values in healthy controls (LOD=-36.7); samples below this threshold were classified as molecularly positive.

### CfMeDIP-seq and Tumor of Origin Prediction

cfMeDIP-seq was performed for each sample as previously described by Shen *et al*. and Wong *et al.* (21,35); See *Supplemental Materials and Methods* detailed immunoprecipitation procedures. CfMeDIP-seq data were processed using the MedRemix pipeline (https://github.com/pughlab/cfMeDIP-seq-analysis-pipeline) as previously described by Wong *et al*. (21), including UMI extraction, alignment to GrCh38, and estimation of bin-wise methylation probabilities using a two-component mixture model.

Following Wong *et al.* (22), we constructed breast- and ovarian-specific classifiers using differentially methylated regions (DMRs) identified by comparing each cancer type against all other samples. The top 150-hyper- and 150 hypomethylated bins from repeated 10-fold cross-validation were aggregated to derive stable, cancer-associated methylation signatures. Average methylation probabilities across these signatures were used as breast and ovarian cancer scores, with higher scores indicating increased likelihood of cancer (see *Supplemental Methods and Materials* for additional details).

### Machine Learning Classification

Supervised models were on trained on paired TS and sWGS samples to classify: (i) cancer-negative *BRCA1/2-carriers* (n=87) versus healthy controls (n=45); and (ii) cancer-positive carriers (n=83) versus cancer-negative carriers with no future cancer diagnosis (n=30), based on clinical follow-up. Input features included six cfDNA-derived scores: FRS, NPS, FS, CGS, TF, and mutation VAF. TF was log1p-transformed; FRS was transformed as -log1p(1 - value); controls were assigned VAF=0.

Four algorithms were evaluated in R using *caret* (v7.0.1) (36): *glm, knn, svmRadial,* and *rf*. Models were trained over 30 random 80/20 train–test splits using four-fold cross-validation (10 repeats), with internal downsampling and normalization (Yeo-Johnson, centering, scaling) applied within each fold. Performance was averaged across splits using AUC, accuracy, precision, recall, F1-score, Kappa, specificity, and 95% confidence intervals.

For task (ii), the top-performing model (by test accuracy) was applied to a held-out validation set of 30 cancer-negative carriers who remained cancer-free (true negative) and 40 who later developed cancer (true positives), using a threshold optimized on the training set via Youden’s Index.

### ctDNA Integration Score

The integration score combined six cfDNA-derived features: FS, NPS, FRS, TF, CGS, and somatic mutation VAF. Only samples with both sWGS and TS data were included; those with unknown diagnoses were excluded, and cancer-negative *BRCA1/2-carriers* diagnosed within 12 months were reclassified as cancer-positive. TF values were log1p-transformed; FRS was transformed as -log1p(1 - value).

Logistic Regression (*glm*) was implemented in R using *caret* (v7.0.1) (36) with 10-fold cross-validation within an 80/20 training-validation split, repeated over 100 iterations with class balancing by downsampling. Final scores were defined as the mean predicted probability across validation sets, using a 0.5 threshold for molecularly positive.

### Healthy Control Cohorts

In total, 45 healthy blood control samples (“Healthy Controls”) were recruited under institutional approval for two cohorts: CHARM (n=35, including 9 individuals with serial blood samples; REB# 19-6239) and Hepatocellular Carcinoma (HCC) healthy controls (n=10; CAPCR# 08-0697). 30 CHARM samples were sequenced with cfMeDIP-seq and included in the methylome analysis. All samples were aligned to GRCh38 and processed according to GATK best practices. Blood processing and computational analyses were performed as described above. See *Supplemental Materials and Methods* for Additional Details.

### Statistical Analysis and Software

All statistical analysis and visualizations were performed using R (v4.3.3, RRID:SCR_001905) in RStudio (2023.12.1.402, RRID:SCR_000432). Statistical significance of cancer scores were calculated using a two-sided Mann-Whitney U test (Wilcoxon rank-sum test).

## Data Availability

All sequencing data files (TS, sWGS, cfMeDIP) are currently being deposited in the European Genome-phenome Archive (EGA) under the accession number EGAS00001006539. Code to reproduce all analyses and figures are available on GitHub (https://github.com/pughlab/EarlyDetectionHBOC). Summary-level data and per-figure source data are provided as Supplementary Tables and in the Supplementary Information. Any additional materials supporting the findings are available from the corresponding authors upon request.

https://ega-archive.org/studies/EGAS00001006539

## Supplemental Tables

Supplemental Table 1 - Sample Descriptions

Supplemental Table 2 - Panel Mutations

Supplemental Table 3 - IchorCNA Tumor Fractions

Supplemental Table 4 - Breast Cancer DNA Methylation Signature

Supplemental Table 5 - Ovarian Cancer DNA Methylation Signature

Supplemental Table 6 - DNA Extraction Information

Supplemental Table 7 - Panel Design

Supplemental Table 8 - Sequencing Coverages

## Endnotes

## Acknowledgments

This work would not have been possible without the patients and their generous participation in this study. This work was supported by grants from the Terry Fox Research Institute (grant #1081) with funds from the TD Ready Challenge (R.H. Kim and T.J. Pugh), and the McLaughlin Centre at the University of Toronto (R.H. Kim and T.J. Pugh). This study was a collaboration with the CHARM Consortium (https://charmconsortium.ca) and is performed under the auspices of the LIBERATE study (<u>NCT 03702309</u>), which is an institutional liquid biopsy program at the University Health Network supported by the BMO Financial Group Chair in Precision Cancer Genomics (Chair held by Dr. Lillian Siu). E.Ensminger is supported by a Terry Fox Research Institute and Canadian Cancer Society Masters Research Training Award. R.H. Kim is supported by Bhalwani Family Charitable Foundation, Goldie R. Feldman, Karen Green and George Fischer Genomics and Genetics Fund, Lindy Green Family Foundation, FDC Foundation, Shar Foundation, The Devine/Sucharda Charitable Foundation, Leslie E. Born, Hal Jackman Foundation, Nicol Family Foundation, James and Christine Nicol, Janice Fukakusa and Greg Belbeck, Jack and Buschie Kamin Foundation, Marcus Tzaferis, Paul Bronfman Family Foundation, The Honey and Leonard Wolfe Family Charitable Foundation, Arman Alie and Margarette Nory, and The Princess Margaret Cancer Foundation. T.J.P. holds the Canada Research Chair in Translational Genomics and is supported by a Senior Investigator Award from the Ontario Institute for Cancer Research and the Gattuso-Slaight Personalized Cancer Medicine Fund. We thank members of the Pugh and Kim Labs for their extensive peer editing and feedback to improve the manuscript. This study was conducted with the support of the Ontario Institute for Cancer Research Genomics Program (http://genomics.oicr.on.ca) enabled through funding provided by the Government of Ontario. Lastly, we particularly thank all of the Hereditary Breast and Ovarian Cancer family members who contributed samples for the study.

## Author Contributions

EE, PL, MB, RHK, and TJP designed and supervised the study. EE, PL and TJP synthesized and interpreted data, and drafted the manuscript and figures. EE performed genomic and fragment analyses. PL performed methylome analyses. S Prokopec, DW, AD, JPB, AN, NZ provided bioinformatics pipeline development support. JE, and S Pedersen performed sample extractions and management. JS, RHK and MB recruited patients, coordinated specimen collection, and synthesized and interpreted data. ML JS and AAM coordinated clinical data and study management. JS, LO and AAM provided clinical support, clinical follow-up, and clinical data abstraction. CM, AD, and PVH provided and annotated imaging data. AA and ML synthesized and interpreted data. BL, and LH performed, coordinated, and managed sample sequencing and data. All authors read, commented, and participated in manuscript editing.

## Additional Information

### Data and Code Availability

All sequencing data files (TS, sWGS, cfMeDIP) are deposited in the European Genome-phenome Archive (EGA) under the accession number EGAS00001006539. Code to reproduce all analyses and figures are available at https://github.com/pughlab/EarlyDetectionHBOC.

### Abbreviations

Cancer Positive (First Positive): *CP-FP* Cancer Positive (Survivor Yes): *CP-SY* Cancer Negative (Survivor Y): *CN-SY* Cancer Negative (Survivor No): *CN-SN* Tumor Fraction: TF Fragment Length Score: FS Fragment Ratio Score: FRS Nucleosome Peak Score: NPS Consensus Griffin Score: CGS

---

**Supplemental Figure 1:**
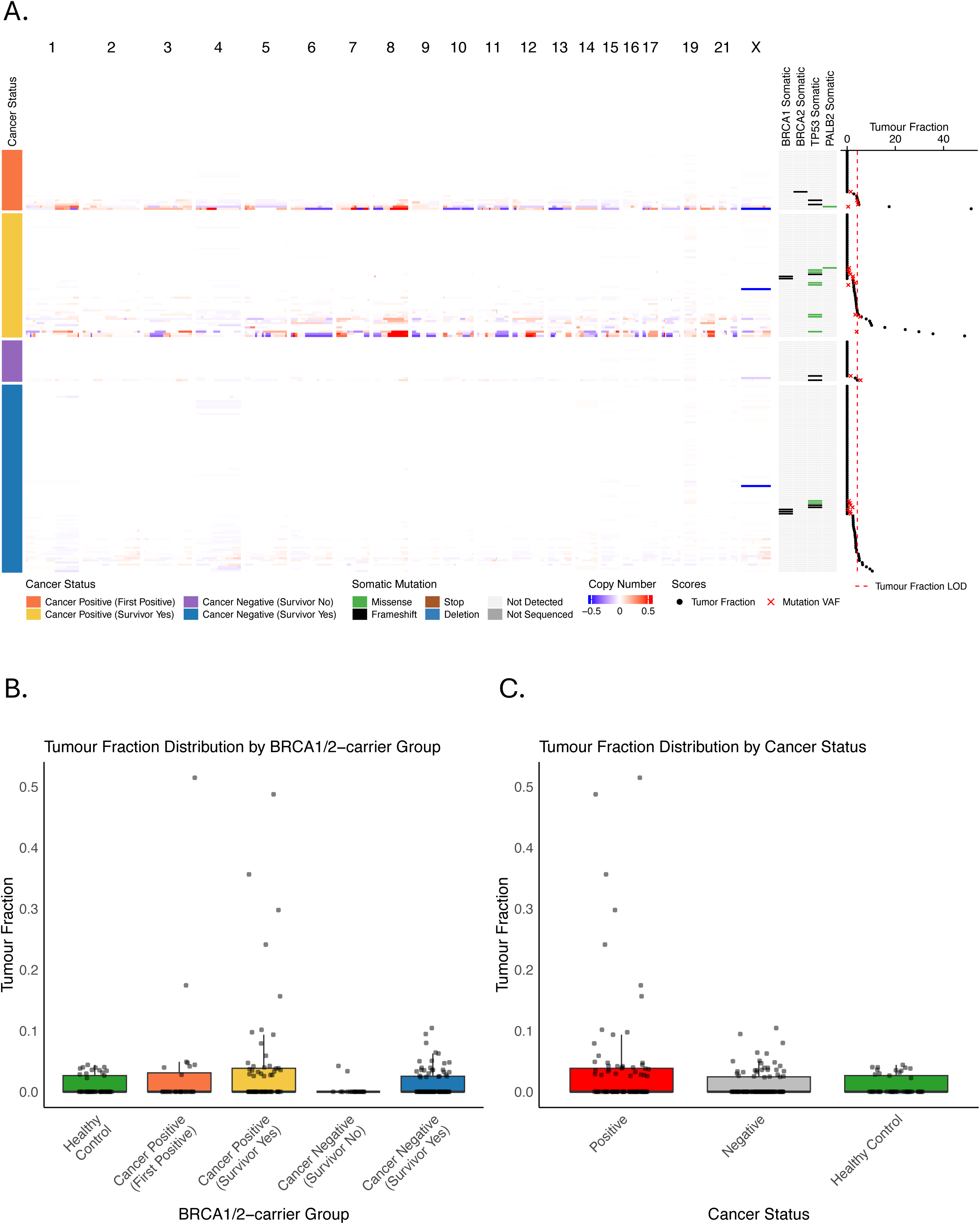
A) Heatmap showing ichorCNA copy number alterations. Somatic mutations from targeted sequencing somatic variant calling analysis and sWGS copy-number informed tumor fractions on the right. Tukey boxplots comparing ichorCNA predicted Tumor Fraction across (B) cancer status groups and healthy control and (C) cancer status and healthy control. P-values were calculated using a two-sided Mann-Whitney U test (Wilcoxon rank-sum test) between all pairwise combinations and adjusted for multiple testing using the Holm method. Only statistically significant comparisons (adjusted p < 0.05) are displayed. * = adj. p < 0.05, ** = adj. p < 0.01, *** = adj. p < 0.001, **** = adj. p < 0.0001.

**Supplemental Figure 2:**
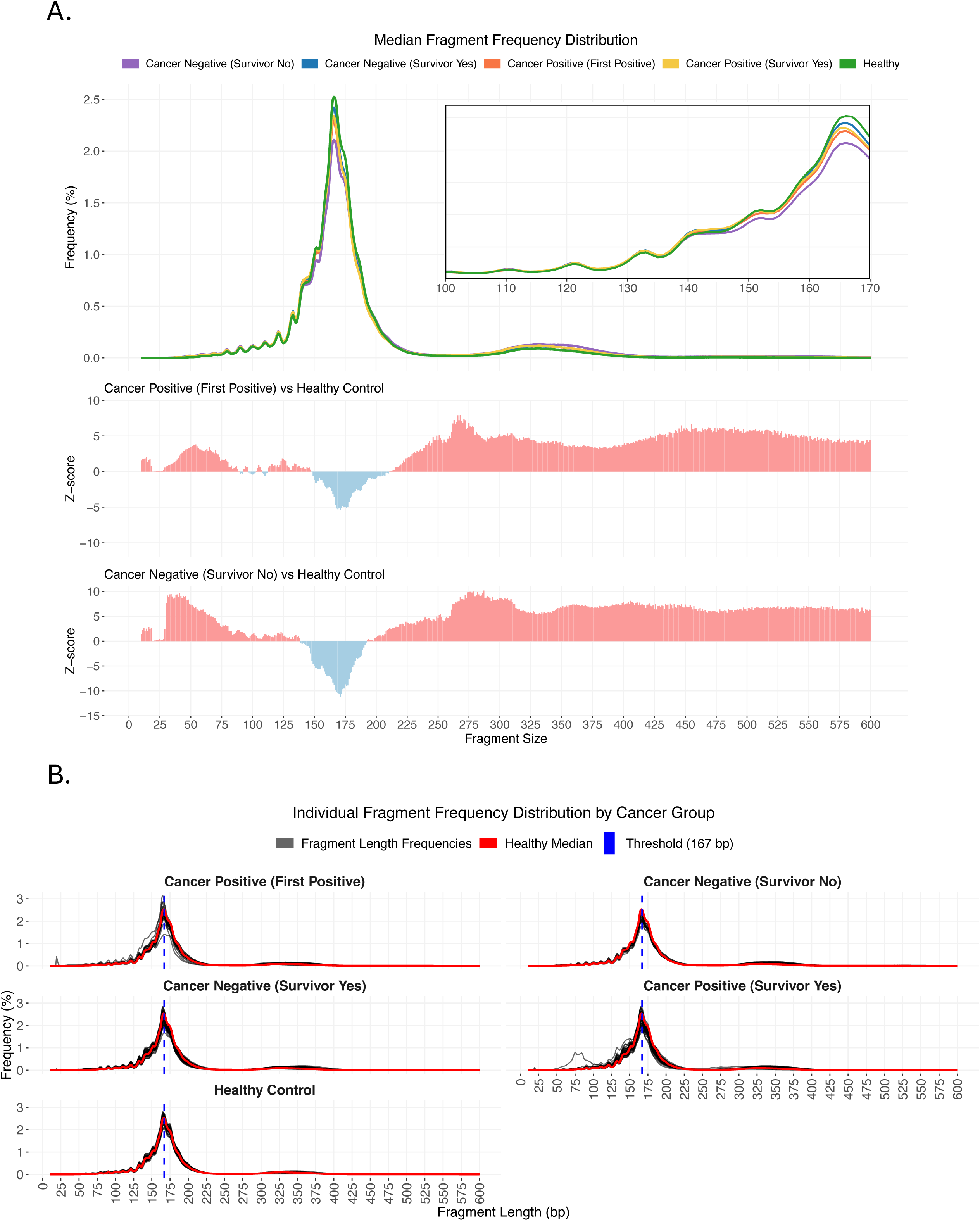
A) Median fragment frequency distribution of samples from healthy controls [green], *BRCA1/2-carrier* Cancer Positive (First Positive) [orange], Cancer Positive (Survivor Yes) [yellow], Cancer Negative (Survivor No) [purple], and Cancer Negative (Survivor Yes) [blue]. Z-scores across the fragment size distribution comparing Cancer Positive (First Positive) to healthy controls (middle) and Cancer Negative (Survivor No) to healthy control (bottom). B) Fragment frequency distributions comparing the *BRCA1/2-carrier* cancer groups and healthy control with the median healthy control (red line).

**Supplemental Figure 3:**
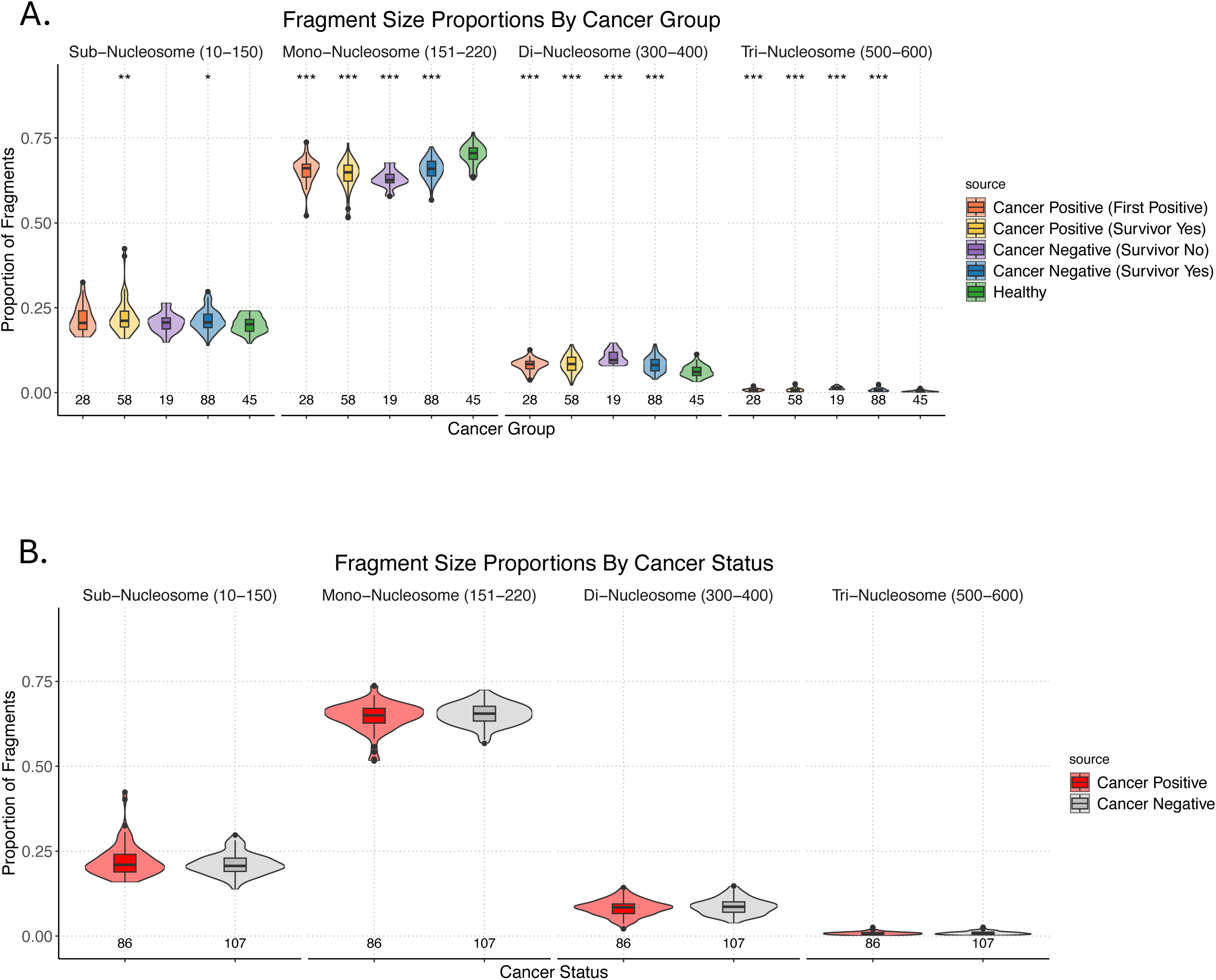
Violin plots showing the fragment size proportions within 4 size compartments: sub-nucleosome (10–150 bp), mono-nucleosomes (151–220 bp), di-nucleosomes (300–400 bp), Tri-nucleosome (500-600 bp) across (A) *BRCA1/2-carrier* groups and healthy control and (C) *BRCA1/2-carrier* cancer positive and negative. *** = p-value < 0.001, ** = p-value < 0.01 * = p-value < 0.05.

**Supplemental Figure 4:**
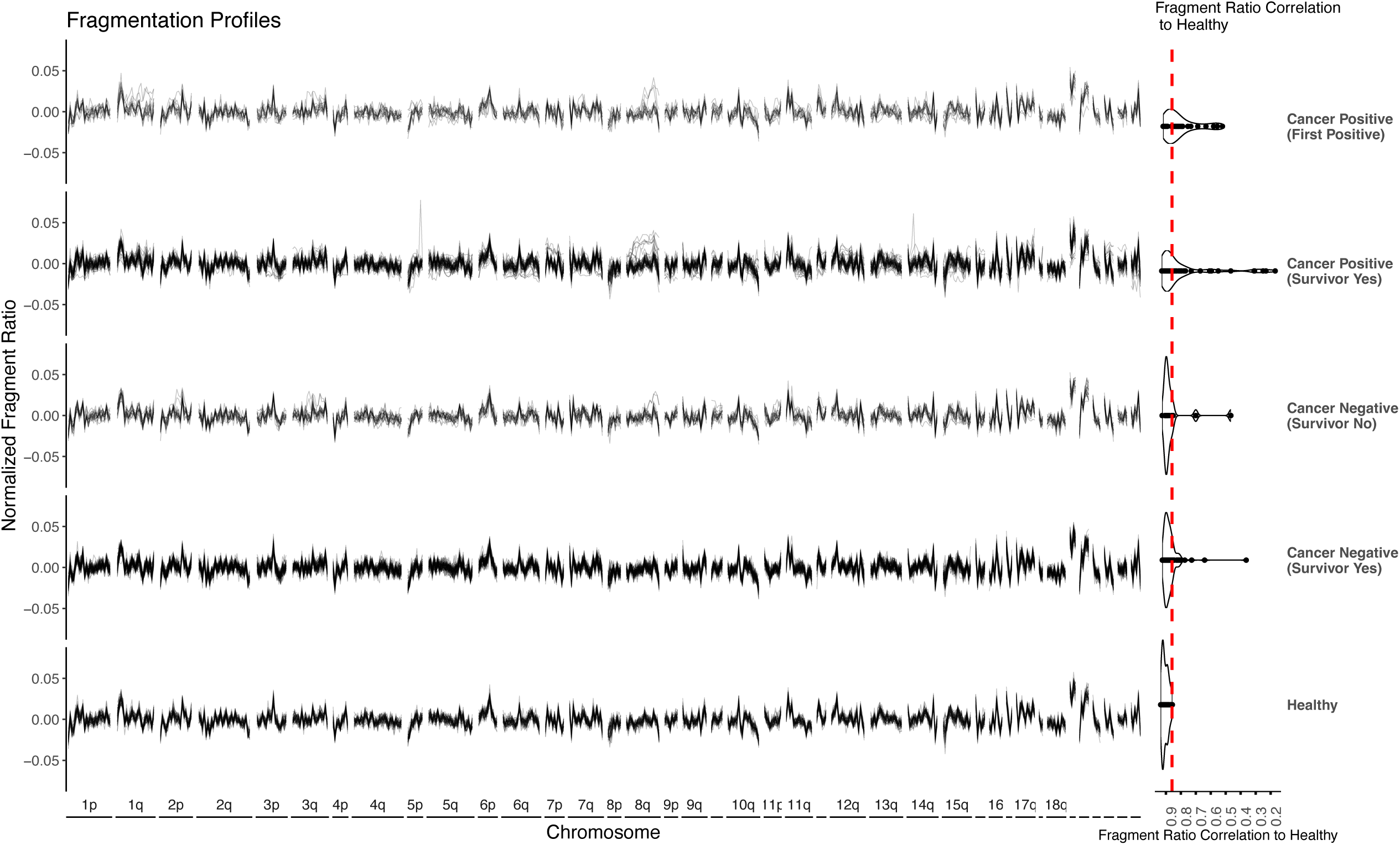
Genome-wide fragment ratio profiles (90-150 bp/151-220 bp) of healthy controls and *BRCA1/2-carriers* separated by cancer status group are shown on the left. Pearson’s correlation scores (Fragment Ratio Score) of *BRCA1/2-carriers* compared to healthy control median shown on the right violin plot; the dashed red line is the Fragment Ratio Score Limit of Detection.

**Supplemental Figure 5:**
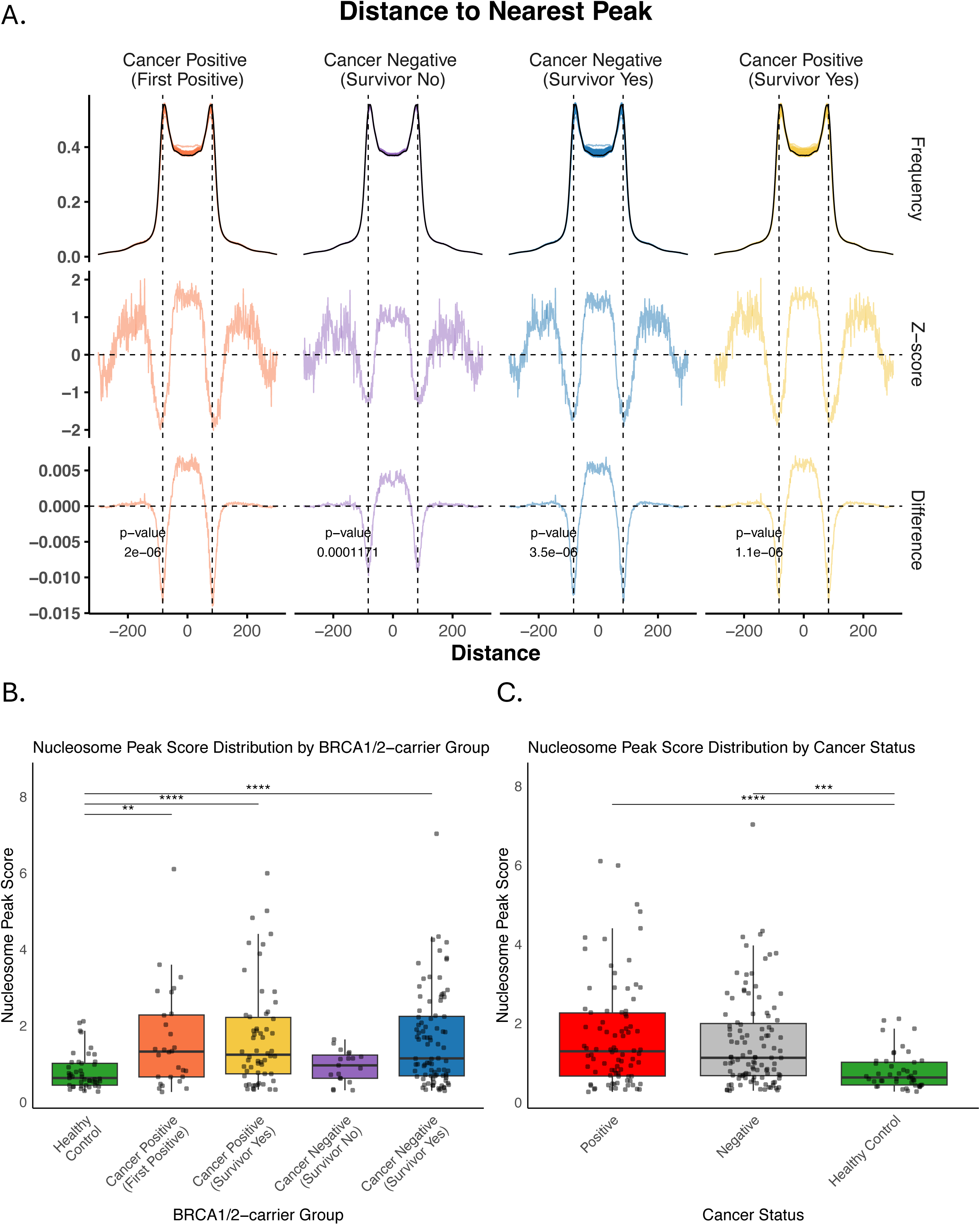
A) Frequency distribution (top), z-scores (middle), and absolute difference (bottom) of the distance of fragment-ends to the closest nucleosome peak in *BRCA1/2-carriers*. Healthy control median is displayed as the black curve in each of the top row panels. p-values were calculated using a two-sided Kolmogorov-Smirnov test. Tukey boxplots comparing Nucleosome Peak Scores across (B) cancer status groups and healthy control and (C) cancer status and healthy control. P-values were calculated using a two-sided Mann-Whitney U test (Wilcoxon rank-sum test) between all pairwise combinations and adjusted for multiple testing using the Holm method. Only statistically significant comparisons (adjusted p < 0.05) are displayed. * = adj. p < 0.05, ** = adj. p < 0.01, *** = adj. p < 0.001, **** = adj. p < 0.0001.

**Supplemental Figure 6:**
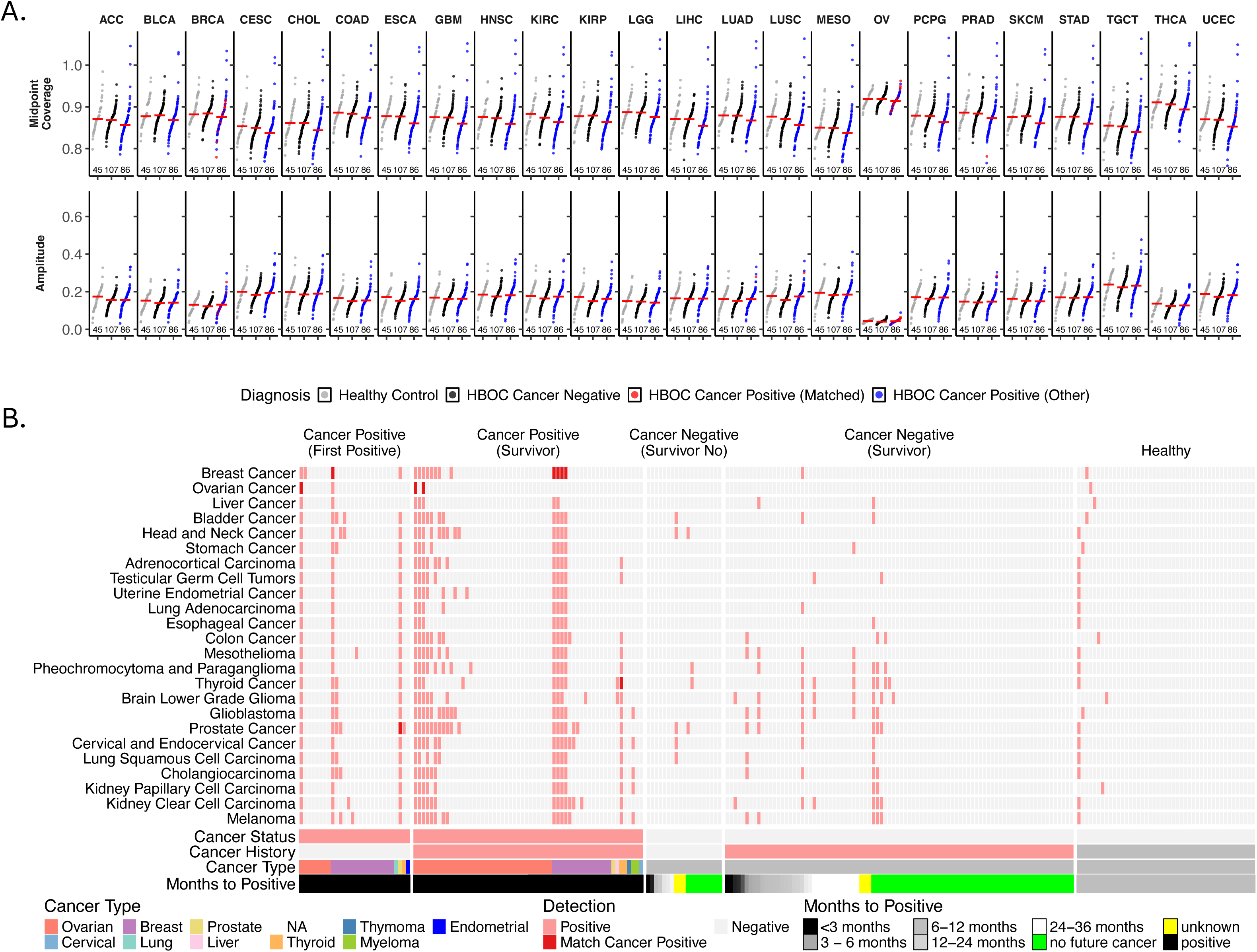
A) Midpoint coverage and amplitude of nucleosome positioning curves of open chromatin sites associated with all TCGA cancer types and one in-house ovarian cancer organoid dataset. P -values calculated using two-sided Student’s t-test. * = p-value < 0.05, ** = p-value < 0.01, *** = p-value < 0.001. B) Heatmap showing the midpoint z-score (relative to healthy control) for all *BRCA1/2-carriers* and healthy control samples with sWGS (columns) across 24 ATAC-seq reference cancer types (rows).

**Supplemental Figure 7:**
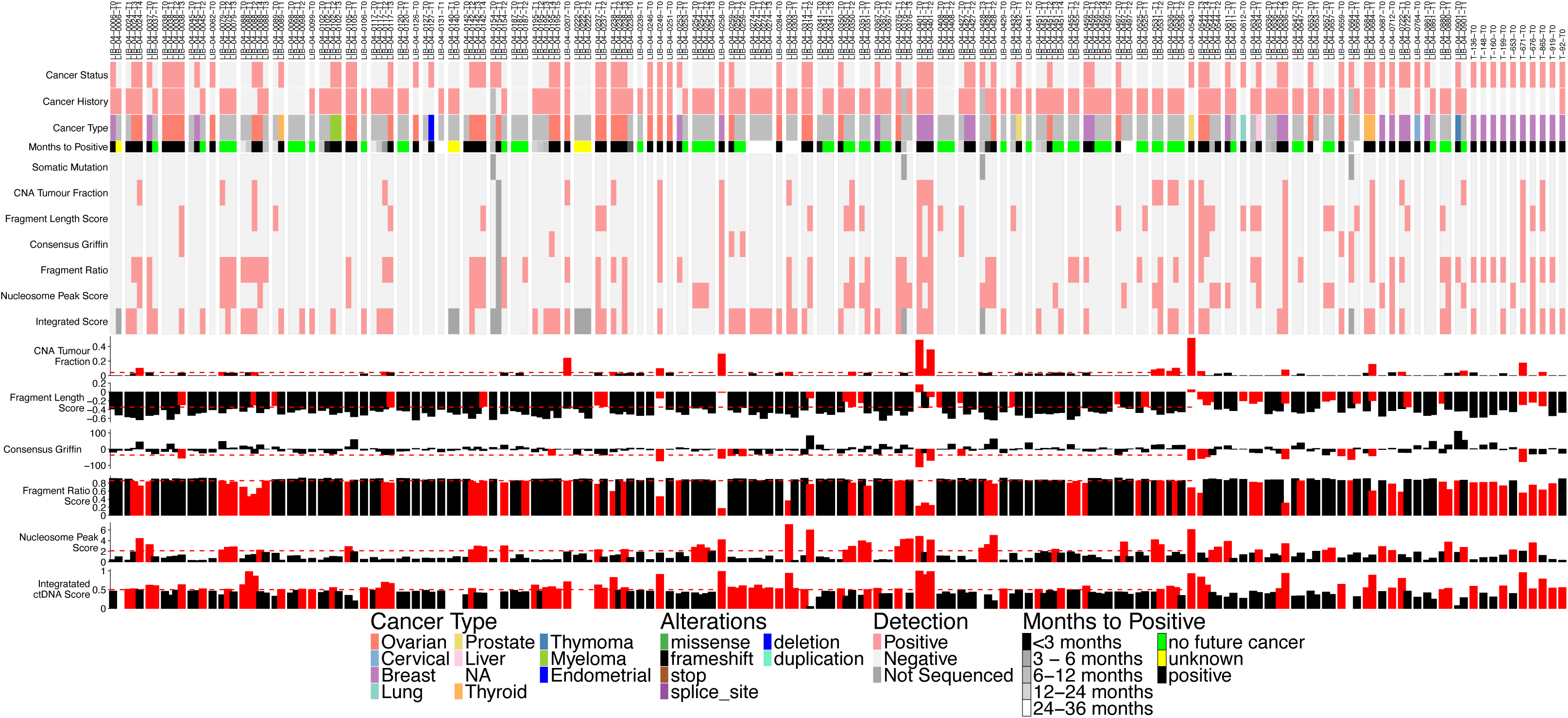
Heatmap and barcharts for each genomic, fragmentomic and integration score, split by patient. Sample and clinical information is displayed at the top (blood sample ID, cancer status, cancer history, cancer type, months to next positive timepoint).

**Supplemental Figure 8:**
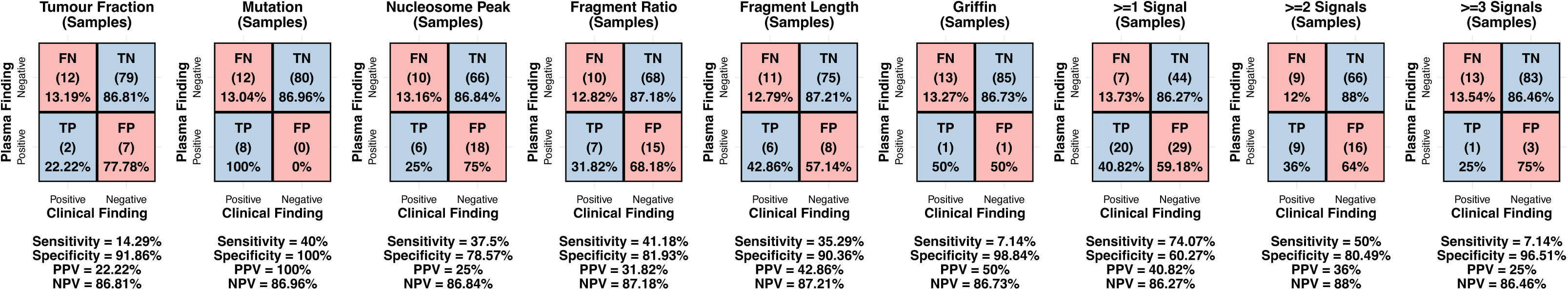
Confusion matrices showing ctDNA and clinical detection rates for Cancer Negative versus *BRCA1/2-carrier* samples for unimodal and integrated analysis. Sensitivity, Specificity, Positive and Negative Predictive Values are listed below each respective plot.

**Supplemental Figure 9:**
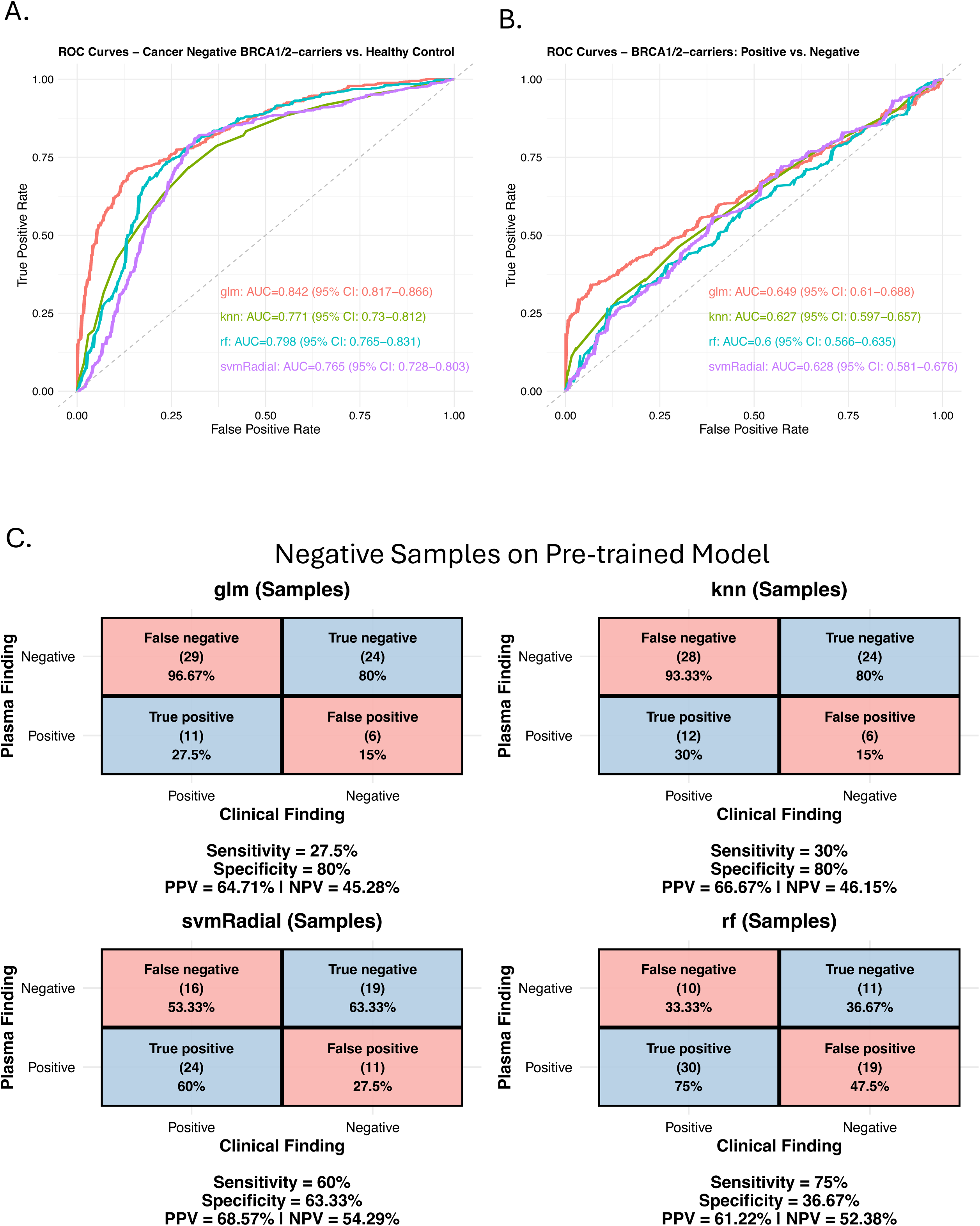
ROC curves for the machine learning model test set of (A) Healthy (Cancer Negative) *BRCA1/2-carriers* versus healthy control and (B) Cancer Positive versus Negative *BRCA1/2-carriers*. AUC values with 95% confidence intervals are displayed and model type being color-matched. C) Confusion matrices showing cancer detection rate for Cancer Negative *BRCA1/2-carrier* samples from pre-trained Cancer Positive versus Negative *BRCA1/2-carriers*. Sensitivity, Specificity, Positive and Negative Predictive Values are listed below each respective plot.

**Supplemental Figure 10:**
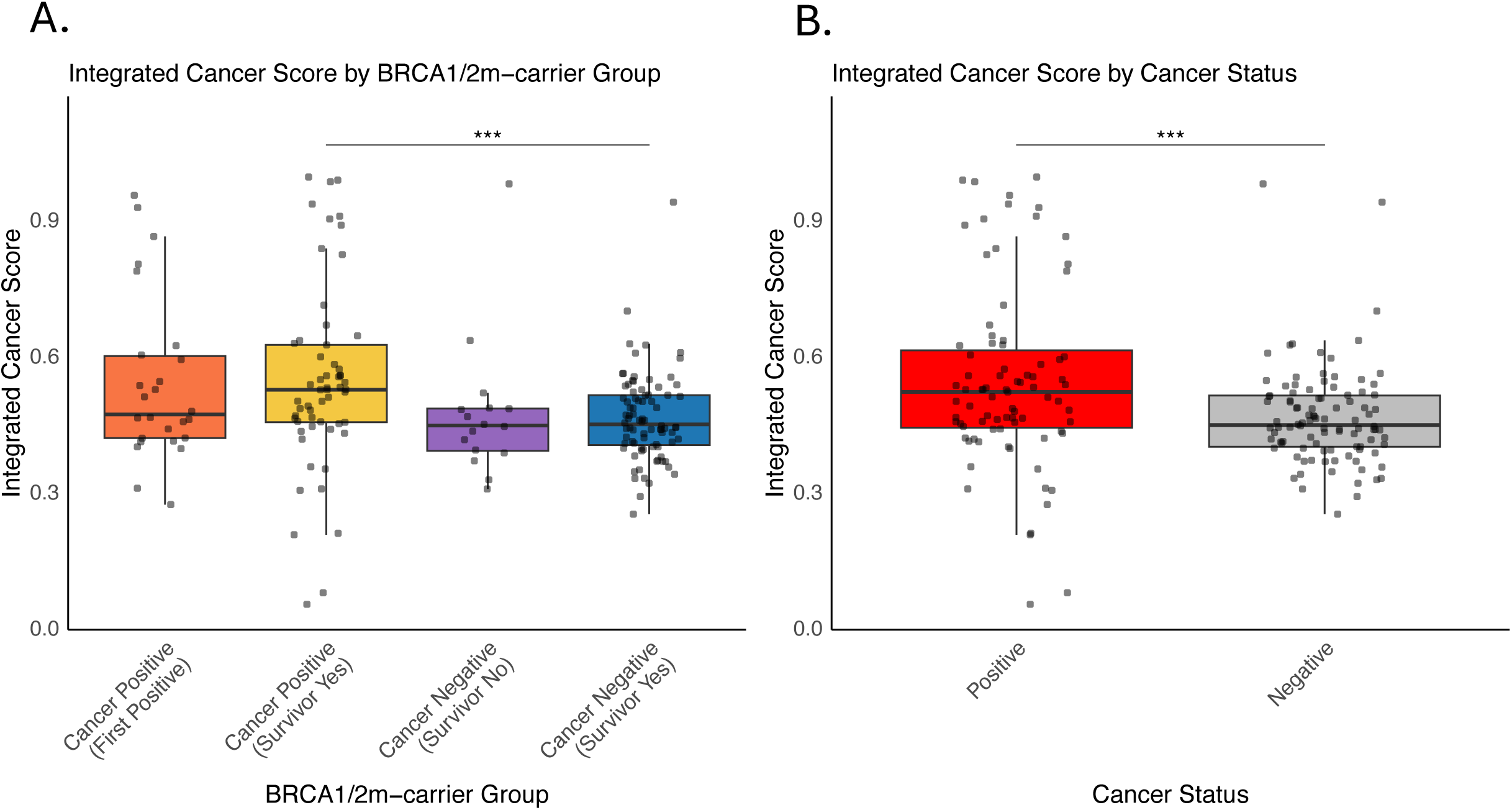
Tukey boxplots comparing Integrated Cancer Score across (A) *BRCA1/2-carrier* groups and (B) cancer status. P-values were calculated using a two-sided Mann-Whitney U test (Wilcoxon rank-sum test) between all pairwise combinations and adjusted for multiple testing using the Holm method. Only statistically significant comparisons (adjusted p < 0.05) are displayed. * = adj. p < 0.05, ** = adj. p < 0.01, *** = adj. p < 0.001, **** = adj. p < 0.0001.

**Supplemental Figure 11:**
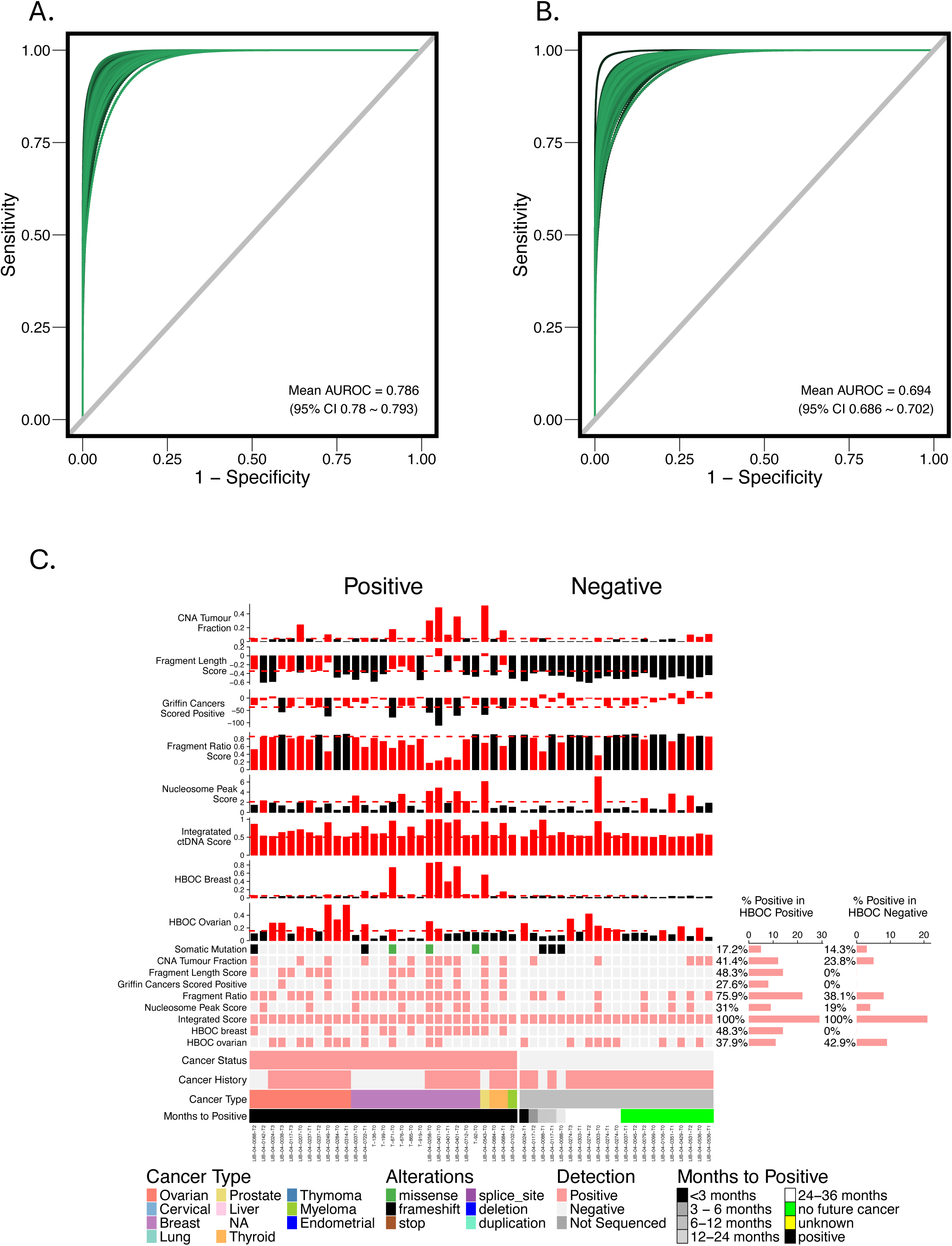
ROC-AUC values for the classification of (A) breast cancer vs other cancers and (B) ovarian cancer vs other cancers using cfMeDIP data (C) Heatmap and barcharts subsetted to samples with positive integrated positive score. Heatmap displays the genomic, fragmentomic, integrated molecular positive scores, and ovarian DNA methylation and breast DNA methylation scores. Clinical information is displayed at the top (cancer status, cancer history, cancer type, months to next positive timepoint). Barchart displaying percentage (%) of cancer-positive and cancer-negative *BRCA1/2-carrier* samples with a molecular positive status for each score are on the right.

## Bibliography

1. Yoshida R. Hereditary breast and ovarian cancer (HBOC): review of its molecular characteristics, screening, treatment, and prognosis. Breast Cancer. 2021;28:1167–80.

2. National Comprehensive Cancer Network: NCCN Clinical Practice Guidelines in Oncology: Genetic/Familial High-Risk Assessment: Breast, Ovarian, Pancreatic and Prostate [Internet]. Report No.: 3.2025. Available from: https://www.nccn.org/professionals/physician_gls/pdf/genetics_bopp.pdf

3. Li S, Silvestri V, Leslie G, Rebbeck TR, Neuhausen SL, Hopper JL, et al. Cancer Risks Associated With BRCA1 and BRCA2 Pathogenic Variants. J Clin Oncol. Wolters Kluwer; 2022;40:1529–41.

4. Gumaste PV, Penn LA, Cymerman RM, Kirchhoff T, Polsky D, McLellan B. Skin cancer risk in BRCA1/2 mutation carriers. Br J Dermatol. 2015;172:1498–506.

5. de Jonge MM, de Kroon CD, Jenner DJ, Oosting J, de Hullu JA, Mourits MJE, et al. Endometrial Cancer Risk in Women With Germline BRCA1 or BRCA2 Mutations: Multicenter Cohort Study. JNCI J Natl Cancer Inst. 2021;113:1203–11.

6. Sherman ME, Foulkes WD. BRCA1/2 and Endometrial Cancer Risk: Implications for Management. JNCI J Natl Cancer Inst. 2021;113:1127–8.

7. National Comprehensive Cancer Network: NCCN Clinical Practice Guidelines in Oncology: Genetic/Familial High-Risk Assessment: Colorectal, Endometrial, and Gastric [Internet]. Report No.: 4.2024. Available from: https://www.nccn.org/professionals/physician_gls/pdf/genetics_ceg.pdf

8. Farncombe KM, Wong D, Norman ML, Oldfield LE, Sobotka JA, Basik M, et al. Current and new frontiers in hereditary cancer surveillance: Opportunities for liquid biopsy. Am J Hum Genet. 2023;110:1616–27.

9. Tung NM, Boughey JC, Pierce LJ, Robson ME, Bedrosian I, Dietz JR, et al. Management of Hereditary Breast Cancer: American Society of Clinical Oncology, American Society for Radiation Oncology, and Society of Surgical Oncology Guideline. J Clin Oncol. Wolters Kluwer; 2020;38:2080–106.

10. Bruhm DC, Vulpescu NA, Foda ZH, Phallen J, Scharpf RB, Velculescu VE. Genomic and fragmentomic landscapes of cell-free DNA for early cancer detection. Nat Rev Cancer. Nature Publishing Group; 2025;1–18.

11. Wan JCM, Massie C, Garcia-Corbacho J, Mouliere F, Brenton JD, Caldas C, et al. Liquid biopsies come of age: towards implementation of circulating tumour DNA. Nat Rev Cancer. Nature Publishing Group; 2017;17:223–38.

12. Kis O, Kaedbey R, Chow S, Danesh A, Dowar M, Li T, et al. Circulating tumour DNA sequence analysis as an alternative to multiple myeloma bone marrow aspirates. Nat Commun. 2017;8:15086.

13. Adalsteinsson VA, Ha G, Freeman SS, Choudhury AD, Stover DG, Parsons HA, et al. Scalable whole-exome sequencing of cell-free DNA reveals high concordance with metastatic tumors. Nat Commun. Nature Publishing Group; 2017;8:1324.

14. Pantel K, Alix-Panabières C. Liquid biopsy and minimal residual disease — latest advances and implications for cure. Nat Rev Clin Oncol. Nature Publishing Group; 2019;16:409–24.

15. Mouliere F, Chandrananda D, Piskorz AM, Moore EK, Morris J, Ahlborn LB, et al. Enhanced detection of circulating tumor DNA by fragment size analysis. Sci Transl Med. 2018;10:eaat4921.

16. Cristiano S, Leal A, Phallen J, Fiksel J, Adleff V, Bruhm DC, et al. Genome-wide cell-free DNA fragmentation in patients with cancer. Nature. 2019;570:385–9.

17. Doebley A-L, Ko M, Liao H, Cruikshank AE, Santos K, Kikawa C, et al. A framework for clinical cancer subtyping from nucleosome profiling of cell-free DNA. Nat Commun. Nature Publishing Group; 2022;13:7475.

18. Shen SY, Burgener JM, Bratman SV, De Carvalho DD. Preparation of cfMeDIP-seq libraries for methylome profiling of plasma cell-free DNA. Nat Protoc. Nature Publishing Group; 2019;14:2749–80.

19. Chan KCA, Jiang P, Chan CWM, Sun K, Wong J, Hui EP, et al. Noninvasive detection of cancer-associated genome-wide hypomethylation and copy number aberrations by plasma DNA bisulfite sequencing. Proc Natl Acad Sci. Proceedings of the National Academy of Sciences; 2013;110:18761–8.

20. Li Y, Xu J, Chen C, Lu Z, Wan D, Li D, et al. Multimodal epigenetic sequencing analysis (MESA) of cell-free DNA for non-invasive colorectal cancer detection. Genome Med. 2024;16:9.

21. Wong D, Luo P, Oldfield LE, Gong H, Brunga L, Rabinowicz R, et al. Early Cancer Detection in Li-Fraumeni Syndrome with Cell-Free DNA. Cancer Discov. 2024;14:104–19.

22. Wong D, Tageldein M, Luo P, Ensminger E, Bruce J, Oldfield L, et al. Cell-free DNA from germline TP53 mutation carriers reflect cancer-like fragmentation patterns. Nat Commun. Nature Publishing Group; 2024;15:7386.

23. Sundby RT, Szymanski JJ, Pan AC, Jones PA, Mahmood SZ, Reid OH, et al. Early Detection of Malignant and Premalignant Peripheral Nerve Tumors Using Cell-Free DNA Fragmentomics. Clin Cancer Res. 2024;30:4363–76.

24. Ongaro G, Petrocchi S, Calvello M, Bonanni B, Feroce I, Pravettoni G. Psychological Determinants of Men’s Adherence to Cascade Screening for BRCA1/2. Curr Oncol. Multidisciplinary Digital Publishing Institute; 2022;29:2490–503.

25. Hesse-Biber S, An C. Within-Gender Differences in Medical Decision Making Among Male Carriers of the BRCA Genetic Mutation for Hereditary Breast Cancer. Am J Mens Health. SAGE Publications Inc; 2017;11:1444–59.

26. Foda ZH, Annapragada AV, Boyapati K, Bruhm DC, Vulpescu NA, Medina JE, et al. Detecting Liver Cancer Using Cell-Free DNA Fragmentomes. Cancer Discov. 2023;13:616–31.

27. Snyder MW, Kircher M, Hill AJ, Daza RM, Shendure J. Cell-free DNA comprises an in vivo nucleosome footprint that informs its tissues-of-origin. Cell. 2016;164:57–68.

28. Vanderstichele A, Busschaert P, Landolfo C, Olbrecht S, Coosemans A, Froyman W, et al. Nucleosome footprinting in plasma cell-free DNA for the pre-surgical diagnosis of ovarian cancer. Npj Genomic Med. Nature Publishing Group; 2022;7:1–9.

29. Ontario Breast Screening Program (OBSP) | Cancer Care Ontario [Internet]. [cited 2025 Jul 4]. Available from: https://www.cancercareontario.ca/en/cancer-care-ontario/programs/screening-programs/ontario-breast-obsp

30. Bao H, Yang S, Chen X, Dong G, Mao Y, Wu S, et al. Early detection of multiple cancer types using multidimensional cell-free DNA fragmentomics. Nat Med. Nature Publishing Group; 2025;1–9.

31. DePristo MA, Banks E, Poplin R, Garimella KV, Maguire JR, Hartl C, et al. A framework for variation discovery and genotyping using next-generation DNA sequencing data. Nat Genet. Nature Publishing Group; 2011;43:491–8.

32. Povysil G, Tzika A, Vogt J, Haunschmid V, Messiaen L, Zschocke J, et al. panelcn.MOPS: Copy-number detection in targeted NGS panel data for clinical diagnostics. Hum Mutat. 2017;38:889–97.

33. Cibulskis K, Lawrence MS, Carter SL, Sivachenko A, Jaffe D, Sougnez C, et al. Sensitive detection of somatic point mutations in impure and heterogeneous cancer samples. Nat Biotechnol. Nature Publishing Group; 2013;31:213–9.

34. Vessies DCL, Schuurbiers MMF, van der Noort V, Schouten I, Linders TC, Lanfermeijer M, et al. Combining variant detection and fragment length analysis improves detection of minimal residual disease in postsurgery circulating tumour DNA of stage II-IIIA NSCLC patients. Mol Oncol. 2022;16:2719–32.

35. Shen SY, Singhania R, Fehringer G, Chakravarthy A, Roehrl MHA, Chadwick D, et al. Sensitive tumour detection and classification using plasma cell-free DNA methylomes. Nature. Nature Publishing Group; 2018;563:579–83.

36. Kuhn M. Building Predictive Models in R Using the caret Package. J Stat Softw. 2008;28:1–26.

