## Supplemental Materials and Methods for "Early Cancer Detection in Hereditary Breast and Ovarian Cancer Syndrome with Cell-Free DNA"

#### Study Design and Patient Cohort

This study was approved by the institutional review boards of the University Health Network (UHN) (REB#: 18-5692), and Jewish General Hospital (JGH) (REB#: MP-05-2020-1928) and conducted in accordance with established ethical guidelines, including the Declaration of Helsinki, CIOMS, the Belmont Report, and the U.S. Common Rule. Written informed consent was obtained from all participants prior to enrolment. All clinical evaluations and treatments were provided by board-certified clinicians according to standard-of-care protocols. Female *BRCA1/2-carriers* follow provincial screening guidelines with annual mammograms and MRIs starting at age 30, clinical breast exams every six months from age 25, and no routine screening for ovarian or pancreatic cancer. However, females with a history of ovarian cancer who had CA-125 as a disease marker may have CA-125 levels monitored every 3–9 months. Male *BRCA1/2-carriers* undergo annual prostate screening from age 40 (PSA and DRE) and clinical breast exams from age 35.

#### Blood Processing and Extraction

Detailed protocols are available at https://charmconsortium.ca/protocols-database/. Venous blood samples were collected in EDTA or Streck tubes (Streck), with EDTA samples processed within 2 hours of collection. Whole blood was centrifuged at 4°C for 10 minutes at 1,900 x g to separate plasma and cellular components. Buffy coats were isolated and stored at −80°C. Plasma was further clarified by a second centrifugation at 4°C for 10 minutes at 16,000 x g to remove residual cells and debris. Purified plasma was stored at −80°C until DNA extraction. See *Supplemental Table 6* for DNA Extraction details.

DNA extraction from plasma was performed using the QIAGEN QIAamp Circulating Nucleic Acid Kit (Qiagen), and genomic DNA from buffy coat was isolated using QIAGEN's AllPrep DNA/RNA/miRNA Universal kit (Qiagen). To ensure compatibility with downstream applications, genomic DNA was fragmented to sizes comparable to cfDNA using an ultrasonicator (LE220, Covaris). Library preparation for TS, WGS and cfMeDIP-seq was conducted only for samples with DNA yields greater than 10 ng. TS was excluded for samples with DNA yields below 40 ng, and samples with less than 10 ng DNA yield were not processed further.

We also addressed several edge cases. Four plasma samples underwent sWGS without matched TS, but corresponding buffy coat TS from other timepoints was available; germline mutations were confirmed using HaplotypeCaller and IGV, and these samples were excluded from somatic analysis. One sample (LIB-04-160) carried a *BRCA1* mutation with an unresolved variant and was classified as *BRCA1*-positive based on buffy data and IGV confirmation. Seven samples from five patients had only gene-level annotations (e.g., *BRCA1* or *BRCA2*) without specific variants; germline status was verified using HaplotypeCaller and IGV. Four samples (LIB-04-0428-T1, LIB-04-0428-T2, LIB-04-0006-T1, and LIB-04-0764-T0) lacked matched buffy coat but had a germline mutation confirmed in plasma TS and manually validated. Similarly, CHARMQ samples (n=7) had only WGS of buffy material, but germline mutations were identified in plasma TS and confirmed by manual review.

#### Cell-Free Methylated DNA Immunoprecipitation

cfMeDIP-seq was performed for each sample as previously described (1,5), using 10 ng of cfDNA libraries tagged with unique molecular identifiers (UMIs), with modifications to the original protocol. Specifically, 0.1 ng of Arabidopsis thaliana methylation control package was added to each 10 ng cfDNA library. This control package contains both methylated and unmethylated spike-in BAC controls (Diagenode). 5% of the sample was aliquoted as a control library, while the remaining portion underwent immunoprecipitation using a monoclonal antibody against 5-methylcytosine (5-mC; Diagenode, clone #33D3, cat. #C15200081-100, RRID:AB_2572207). Both the immunoprecipitated and control libraries were subsequently amplified, indexed, and pooled for sequencing.

#### Copy number alteration and Tumor Fraction Estimation

Generation of an in-house panel of normals (PON; n=45), copy number alteration (CNA) analysis and tumor fraction estimation were performed using ichorCNA (version 0.2.0; Broad Institute; https://github.com/broadinstitute/ichorCNA) as described by Addalsteinsson *et al.* (6). All analyses, including PON generation and sample processing were executed using the PughLab pipeline-suite (https://github.com/pughlab/pipeline-suite). LOD was set to the 99th percentile of TF in healthy controls (0.0424); samples exceeding this threshold were classified as molecularly positive.

#### Fragment Length Analysis and Score Calculation

Global fragment length distributions were computed using Picard CollectInsertSizeMetrics (v4.0.1.2; RRID:SCR_006525), retaining fragment lengths between 1 to 599 bp. Fragment length scores were calculated using a previously published reference set from Vessies *et al.* and described further in Wong *et al.* (1,7). Briefly, the Vessies *et al.* reference set assigns log2 difference-based weights to fragment lengths (1–600 bp) based on differences in fragment frequency distributions between tumor-informed and healthy cfDNA. Sample-level fragment length scores were derived by mapping all observed fragment lengths to this reference and calculating the mean of the assigned weights. The LOD was defined as the 99th percentile of healthy control scores (LOD=-0.346); samples exceeding this threshold were classified as molecularly positive.

#### Fragment Ratio Analysis and Score Calculation

Fragment ratio was based on the work of Cristiano *et al.* (8) DELFI approach for hg38 reference genome and further adapted from Wong *et al.* (9). Briefly, the ratio of short (90–150 bp) to long (151–220 bp) mapped cfDNA fragments was calculated across the genome in non-overlapping 5 Mb bins, excluding ENCODE GRCh38 blacklisted regions. GC bias was corrected using LOESS regression (span=0.75), and bin-wise coverage was computed on the GC-adjusted ratios to reduce technical noise. Fragment Ratio Scores were derived by correlating each bin-wise profile with the healthy control median using Pearson correlation. The LOD was defined as the 1st percentile of healthy control correlations (LOD=0.859), with samples below this threshold classified as molecularly positive.

#### Nucleosome Positioning Analysis and Score Calculation

Nucleosome Positions were analyzed as previously described by Wong *et al.* (9). Briefly, healthy control nucleosome peaks were obtained from Snyder *et al.* (10). Deduplicated bam files were converted into BED files with fragment starts and ends using Bedtools (v2.27.1; RRID:SCR_006646). Fragment starts and ends were then mapped to the closest nucleosome peak and the distance calculated. Only fragments that mapped within 1000 bp from a peak were kept for downstream analysis (11).

Nucleosome peak scores were derived by analyzing cfDNA fragments spanning the nucleosome region (-/+83 bp; ~167 bp). Bin-wise z-scores were calculated based on the mean of healthy controls and averaged to obtain a final score per sample. The LOD was set as the 99th percentile of scores from healthy controls (LOD=2.1), with samples above this threshold classified as molecular positive.

#### Nucleosome Accessibility and Score Calculation

Nucleosome positions were analyzed using Griffin tool (v0.1.0; https://github.com/adoebley/Griffin) as described in Doebley *et al.* (12) using 90-220bp and adapted as previously described by Wong and colleagues (9). We solely used the 10,000 sites from tissue specific open-chromatin sites. In addition to the 23 TCGA references, we generated an in-house custom ovarian ATAC-seq reference of the 10,000 most important sites. Briefly, to generate this in-house reference set, we analyzed chromatin accessibility from 11 HGSOC organoid samples processed using the ATAC-seq protocol as described by Boutzen *et al.* (13). For each sample, peaks were called and stored in standard .narrowPeak format, compatible with MACS2 output. To quantify chromatin accessibility across samples, a non-overlapping union peak set was first generated by merging peaks from all HGSOC organoid samples into a consensus peak file. Each sample's .narrowPeak file was then intersected with the consensus set using bedtools (v2.30.0; RRID:SCR_006646), counting the number of overlapping peaks per region. This produced a matrix of accessibility counts across all 11 samples. Row-wise sums were computed, and the 10,000 most accessible consensus regions were selected. These top-ranked regions were saved as a BED-format file containing genomic coordinates and peak metadata, and were integrated into the in-house pipeline-suite (https://github.com/pughlab/pipeline-suite).

To compute the Consensus Griffin Score (CGS), we calculated z-scores of fragment coverage at five midpoint positions (−30, −15, 0, 15, 30) for each cancer type, relative to the healthy control median. All z-scores across the 24 cancer types were then summed to yield a single CGS per sample. The limit of detection (LOD) was defined as the 99th percentile of CGS values in healthy controls(LOD=-36.7); samples below this threshold were classified as molecularly positive.

### Tumor of Origin Detection Using cfMeDIP-seq

The raw cfMeDIP-seq FASTQ files were processed using the MedRemix pipeline (https://github.com/pughlab/cfMeDIP-seq-analysis-pipeline) (14). Initially, UMI barcodes were extracted and spacers removed using the extract_barcodes.py script from the ConsensusCruncher toolkit (https://github.com/pughlab/ConsensusCruncher). Reads were then aligned to the human reference genome (GRCh38/hg38) using BWA-MEM (v0.7.17; RRID:SCR_010910), followed by sorting and indexing with Samtools (v1.14; RRID:SCR_002105) . For downstream quantification, paired-end reads were summarized across non-overlapping 300 bp genomic windows (bin), and CpG counts were recorded per window. Coverage data were modeled using a two-component mixture model to distinguish between methylated and unmethylated regions, incorporating CpG density and GC content as covariates through negative binomial regression. The unmethylated component was estimated from windows lacking CpGs, with coverage modeled as a function of GC content. Methylation probabilities were then inferred for each genomic window through an expectation-maximization algorithm, estimating both mixing proportions and regression parameters until convergence.

Following the generation of bin-wise methylation probabilities, cancer-type-specific classifiers were constructed to distinguish breast and ovarian cancers from other samples in the cohort. To identify informative features, DMRs were determined by comparing methylation profiles of breast (ovarian) cancer samples against all other samples using the limma R package (v3.56.2; RRID:SCR_010943). The top 150 hypermethylated and 150 hypomethylated bins were selected as candidate DMRs for classifier training. A robust evaluation strategy was employed using 10-fold cross-validation repeated 100 times to reduce the impact of random split. Across these iterations, the top 150 hypermethylated regions identified in each limma comparison were aggregated, and regions consistently selected in all 100 runs were retained as stable, cancer-associated methylation signatures. These refined signatures were then treated as breast (ovarian) signatures, and the average methylation probability of them was used as the breast (ovarian) scores. The higher the score, the more likely the sample is cancer positive.

### Machine Learning Classification

We trained supervised machine learning models on paired targeted sequencing (TS) and shallow whole-genome sequencing (sWGS) samples to classify (i) cancer-negative BRCA1/2-mutation carriers (n=87) versus healthy controls (n=45), and (ii) cancer-positive carriers (n=83) versus cancer-negative carriers with no future cancer diagnosis (n=30), based on clinical follow-up data. Input features included six cfDNA-derived scores: fragment ratio, nucleosome peak, fragment size (FS), Griffin consensus, tumor fraction (TF), and mutation variant allele fraction (VAF). TF were log1p-transformed; FRS was transformed as −log1p(1 − value); mutation VAF of controls was assumed to be 0.

We evaluated five machine learning algorithms: logistic regression (glm), k-nearest neighbors (knn), support vector machines (svmRadial), random forest (rf). Models were trained over 30 random 80/20 train–test splits using four-fold cross-validation, 10 repeats with *caret* (v7.0.1)

(15), applying internal downsampling and normalization (Yeo-Johnson, centering, scaling) within each fold. Generalization performance was estimated by averaging validation metrics across iterations, including AUC, accuracy, precision, recall, F1-score, Kappa, specificity, and 95% confidence intervals. For task (ii), the best-performing model based on test accuracy was applied to a held-out validation set comprising 30 cancer-negative carriers who remained cancer-free (true negatives) and 40 who later developed cancer (true positives), using a threshold optimized on the training set via Youden's Index.

#### ctDNA Integration Score

The ctDNA integration score was derived from six features: FS, nucleosome peak score, fragment ratio score, TF, consensus griffin score, and mutation status. Only samples with both sWGS and TS data were included; those with unknown diagnoses were excluded, and cancer-negative BRCA1/2-carriers diagnosed within 12 months were reclassified as cancer-positive. TF values were log1p-transformed; FRS was transformed as -log1p(1 - value). Class sizes were balanced by downsampling in each iteration. Logistic regression (glm) was performed using caret R package (v7.0.1) (15) with 10-fold cross-validation within an 80/20 training–validation split, repeated over 100 iterations. Final scores were defined as the mean predicted probability across validation sets, with a 0.5 threshold for molecular positivity.

#### Healthy Control Cohorts

In total, 45 healthy blood control samples ("Healthy Controls") were recruited under institutional approval for two cohorts: CHARM (n=35, including 9 individuals with serial blood samples; REB# 19-6239) and Hepatocellular Carcinoma (HCC) healthy controls (n=10; CAPCR# 08-0697). 30 CHARM samples were sequenced with cfMeDIP-seq and included in the methylome analysis. All samples were aligned to GRCh38 and processed according to GATK best practices. Blood processing and computational analyses were performed as described above. Eligible participants were adults who provided written informed consent, no personal history of cancer, autoimmune disease, or chronic inflammation and had not been diagnosed with hereditary cancer syndrome. Individuals unable to provide informed consent or who were pregnant were excluded. HCC healthy controls were recruited as living donors of liver tissue and went through extensive screening for cancer (16).

12. Doebley A-L, Ko M, Liao H, Cruikshank AE, Santos K, Kikawa C, et al. A framework for clinical cancer subtyping from nucleosome profiling of cell-free DNA. Nat Commun. Nature

Publishing Group; 2022;13:7475.

13. Boutzen H, Madani Tonekaboni SA, Chan-Seng-Yue M, Murison A, Takayama N, Mbong N, et al. A primary hierarchically organized patient-derived model enables in depth interrogation of stemness driven by the coding and non-coding genome. Leukemia. 2022;36:2690–704.

14. Stutheit-Zhao EY, Sanz-Garcia E, Liu Z (Amy), Wong D, Marsh K, Abdul Razak AR, et al. Early Changes in Tumor-Naive Cell-Free Methylomes and Fragmentomes Predict Outcomes in Pembrolizumab-Treated Solid Tumors. Cancer Discov. 2024;14:1048–63.

15. Kuhn M. Building Predictive Models in R Using the caret Package. J Stat Softw. 2008;28:1–26.

16. Chen K, Li Z, Kirsh BO, Luo P, Pedersen S, Bucur RC, et al. Plasma cell-free DNA methylomes for hepatocellular carcinoma detection and monitoring after liver resection or transplantation [Internet]. medRxiv; 2024 [cited 2025 July 31]. page 2024.10.01.24314116. Available from: https://www.medrxiv.org/content/10.1101/2024.10.01.24314116v1
